# Predicting severe COVID-19 outcomes for triage and resource allocation

**DOI:** 10.1101/2021.04.12.21255201

**Authors:** Anthony Onde Morada, Caleb Scheidel, Jennifer Brown, Jeremy Albright, Victor Kolade, Burt Cagir

**Affiliations:** Guthrie Robert Packer Hospital, Sayre, Pennsylvania, United States of America; Geisinger Commonwealth School of Medicine, Scranton, PA, United States of America; Methods Consultants, Ypsilanti, Michigan, United States of America

## Abstract

**Background:** While numerous studies have identified factors associated with severe COVID-19 outcomes, they have yet to quantify these characteristics. Therefore, our study’s purpose is to stratify these risk factors and use them to predict outcomes.

**Study Design:** This is a retrospective review of the CDC COVID-19 Surveillance Data. Logistic regression models calculated risk estimates for independent variables, and random forest models predicted the chance of severe outcomes.

**Results:** Our sample of 3,798,261 patients with COVID-19 consisted mainly of females (51.9%), 10-to 69-year-olds, and White/Non-Hispanics (34.9%). Most were not healthcare workers (90.6%) and did not have preexisting medical conditions (47.1%). Age had an increased risk of severe outcomes that grew every decade of life. White patients had a decreased occurrence of severe outcomes than Non-Whites, except for Pacific Islanders with comparable mortality. The variable selection algorithm detected that three outcomes were more accurate without healthcare worker classification: mechanical ventilation/intubation, pneumonia, and ARDS Acute respiratory distress. However, providers had a decreased risk of severe outcomes overall. Also, patients with preexisting conditions demonstrated an increased risk in all outcomes. Compared to the logistic regressions, the predictive models had a higher performance (AUC>0.8). The death model had the best metrics, followed by hospitalization and ventilation. We amassed these predictive models into the Severe COVID-19 Calculator web application that estimates the probability of severe outcomes.

**Conclusions:** Several patient social and medical demographics recorded by the CDC significantly affect severe COVID-19 outcomes suggesting a multifactorial influence. To account for these variables, a generated Severe Covid-19 Calculator can accurately predict the chance of severe outcomes in citizens that may contract or have COVID-19.

## Introduction

As the U.S. transitions into the vaccine era of the coronavirus disease-19 (COVID-19) pandemic, health care providers and administrators have to plan allocation of COVID-19 treatments and reconsider reopening elective hospital services and surgeries. Therefore, hospitalists and surgeons must balance a patient’s risk of nontreatment against potential hospital-acquired COVID-19 infection as hospitals return to pre-pandemic volumes [1–3]. Simultaneously, the government charged hospital administrators and local health care leaders of each state, tribe, and territory to develop their vaccine distribution plan [4–6]. Since the U.S. Food and Drug Administration (FDA) granted emergency use authorization of the Moderna and Pfizer-BioNTech COVID vaccines in late December 2020, the U.S. continues to trail other countries in the proportion of citizens that received at least one vaccine dose [4–9].

While investigators and journalists have yet to agree, most experts have suggested that technical challenges, lack of federal involvement, and strict adherence to state and CDC priority groups have contributed to the vaccine distribution and administration gap – on January 6th, this gap was as large as 3.7 million to 603,000. [10] Others attribute this vaccination gap to confusing and ambiguous state guidance or poor distribution infrastructure [11–13]. Part of this problem stems from a long-standing ethical question in medicine: How do we distribute a limited treatment equitably and fairly? The diverse array of COVID-19 symptoms complicates this question as severe outcomes, such as hospitalization and death, occur more often in select subgroups, while others are asymptomatic. Researchers also discovered that not unlike our healthcare system pre-pandemic, the social determinants of health amplified these deadly outcomes [14]. A report from the Proceedings of the National Academy of Sciences (PNAS) of the United States of America demonstrated that Black and Latino Americans would be disproportionally left out the CDC Phase 1b recommendation of persons ≥75 years of age as minority groups die younger than their white counterparts [15,16].

While numerous studies have qualified risk factors associated with severe COVID-19 outcomes, the current literature has yet to quantify these objective characteristics. Therefore, the purpose of this study is to determine which citizens would most likely develop severe COVID-19 if they contracted the disease. Similar to how the Model for End-Stage Liver Disease (MELD) aids the distribution of livers, our goal is to create an objective and multifactorial algorithm that can stratify patients at risk for severe COVID-19 outcomes [17]. To fulfill these goals, we conducted a retrospective review of the CDC COVID-19 Case Surveillance Restricted Access Detailed Data to enumerate severe COVID-19 outcomes and create a prediction algorithm with machine learning that stratifies a citizen’s risk for severe COVID-19 outcomes.

## Methods

### Data Collection

To determine the rates of severe COVID-19 outcomes, we obtained the CDC COVID-19 Case Surveillance Restricted Access Detailed Data after completion and approval through the Registration Information and Data Use Restrictions Agreement (RIDURA) and restricted data access process [18]. The six severe COVID-19 outcomes recorded in the CDC data include hospitalization, intensive care unit (ICU) admission, mechanical ventilation or intubation, pneumonia, acute respiratory distress syndrome (ARDS), and death.

To ensure our sample best represents the COVID-19 population, we only included patients with laboratory-confirmed COVID-19, thereby excluding suspected cases. Additionally, we chose a study cut-off date of December 15th, 2020, which corresponds to the first day of vaccination in the U.S., to eliminate any potential vaccination effects. Therefore, only positive cases with a date of first specimen collection before December 15th, 2020, were included in our study. We also converted the CDC state and region data into the Census Bureau Regions, which reclassifies each patient’s residence into one of the five regions: The Northeastern, Midwestern, Southern, Western, and U.S. Territory Regions to account for potential geographic confounding [19].

The information presented in the database reflects the CDC Human Infection with 2019 Novel Coronavirus Person Under Investigation (PUI) and Case Report Forms at an individual patient level. The variable “pre-existing medical conditions” also reflects this reporting form and is defined as having any of the following conditions: chronic lung disease, diabetes mellitus, cardiovascular disease, chronic renal disease, chronic liver disease, immunocompromised condition, neurologic/neurodevelopmental/intellectual disability, other current diseases, current pregnancy status, and current or former smoker [18].

The CDC Surveillance Review and Response Group (SRRG), as part of the CDC’s COVID-19 Emergency Response, maintains the COVID-19 Case Surveillance Restricted Access Detailed Dataset. While the CDC approves database access and study proposals through evaluation of the RIDURA. Institutional review board approval was not required as this study involves data that is a collection of publicly available data with information recorded by the SRRG in such a manner that the subjects cannot be identified directly or through identifiers linked to the subjects (45 Code of Federal Regulations (CFR) 46.101(b)). The CDC does not take responsibility for the scientific validity or accuracy of methodology, results, statistical analyses, or conclusions presented.

### Determining Risk Factors

Baseline demographic variables and clinical outcomes were presented with frequencies and percentages. The following outcomes were considered: hospitalization, ICU admission, mechanical ventilation/intubation, pneumonia, acute respiratory distress, and mortality. For each outcome, an initial multivariable logistic regression model fit all baseline demographics and symptoms as covariates. Forest plots showing odds ratios and 95% confidence intervals for each covariate in the model were presented, and model fit metrics including Akaike information criterion (AIC), Bayesian information criterion (BIC), Area under the curve (AUC), and pseudo-R2 measures were shown. The Least Absolute Shrinkage and Selection Operator (LASSO) logistic regression was then used as a variable selection technique to assess if any variables were uninformative to predicting each outcome. The LASSO algorithm’s penalty hyperparameter was tuned using 5-fold cross-validation to determine its optimal value based on the area under the receiver operating characteristic curve (AUC). Variables that were shrunk to zero in the LASSO models were dropped, and another logistic regression model was fit for each outcome with the variables kept by the LASSO model. Model fit metrics were then presented for these models to determine if dropping uninformative variables improved model fit. Statistical significance was defined as a p-value < 0.05.

### Predictive Model Creation

For each outcome, a random forest model was fit with baseline demographics, including age, sex, race/ethnicity, region, healthcare worker, pre-existing medical conditions, and month of positive COVID-19 test as predictive features.

Before modeling, the data was first split into training and testing sets. Due to the large sample size, we took a random sample of 3% of the data for computational speed. We then took a random sample of 80% of that data to train the model and used the remaining 20% of the data as the test set. Data pre-processing included creating indicator variables for all non-numeric categorical values of the predictor variables and using indicators of the month of the positive test date for accounting treatment improvement over time. Since fewer patients experienced each outcome than those who didn’t, we upsampled each model’s outcome to balance the classes.

The random forest algorithm’s hyperparameters were tuned using 3-fold cross-validation on the training set, with the number of trees contained in the ensemble fixed at N = 1000. The hyperparameters correspond to the number of predictors randomly sampled at each split when creating the tree models (m_try) and the minimum number of data points in a node required for the node to be split further (min_n). The optimal hyperparameters were chosen based on which values maximized the area under the receiver operating characteristic curve (AUC). The AUC metric measures fit from 0.5 (no better than a coin flip) to 1.0 (perfect prediction).

Final models were fit on the entire training set and evaluated on the test set using the following metrics to assess performance: accuracy, precision, recall, and AUC. Accuracy is the number of correct predictions divided by the total number of predictions or the percentage of predictions from the correct model. Precision is the number of true positives divided by the sum of the true and false positives, demonstrating how precise a model is out of those it predicted positive. The recall is the number of true positives divided by the true positives and false negatives, accounting for the percentage of relevant results correctly classified by the model.

The random forest models’ relative predictive performance was compared to the logistic models’ performance in risk factor estimation. Feature importance plots for the random forest models were produced to show which variables were most important to making the predictions for each outcome. All analyses were performed in R Version 4.0.3 [20].

## Results

The total sample contained 3,798,261 patients with laboratory-confirmed cases of COVID-19 between January 1st, 2020, and December 15th, 2020. Table1 presents the sample demographics. Our sample of patients with COVID-19 were primarily women between the ages of 10 - 69, white/non-Hispanic, and who lived in the Northeast CDC Census Regions. While most patients’ status regarding healthcare work or pre-existing medical conditions was missing or unknown, most subjects with COVID-19 were not healthcare workers and did not have any pre-existing medical conditions (Table 1).

**Table 1.**
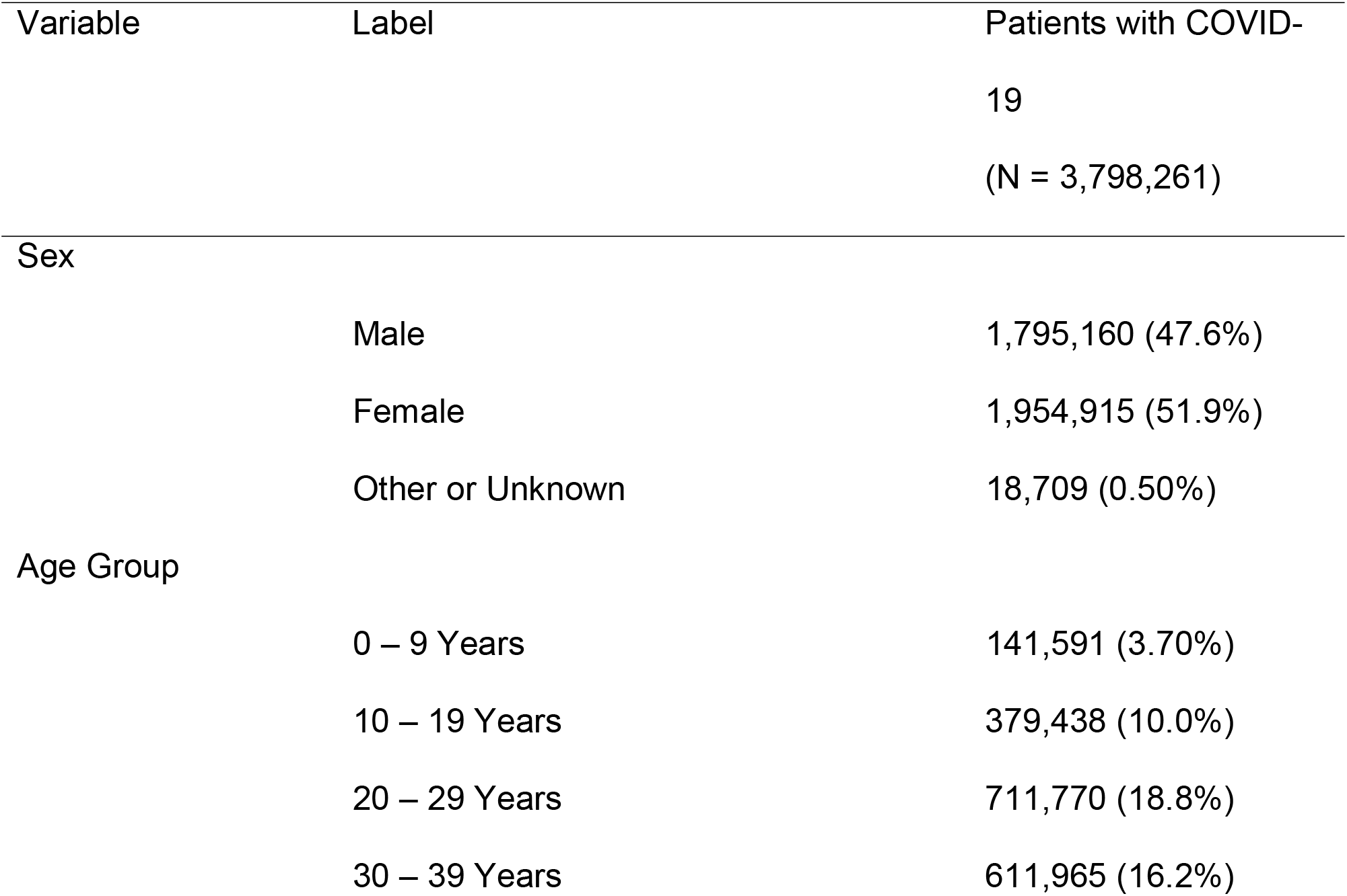
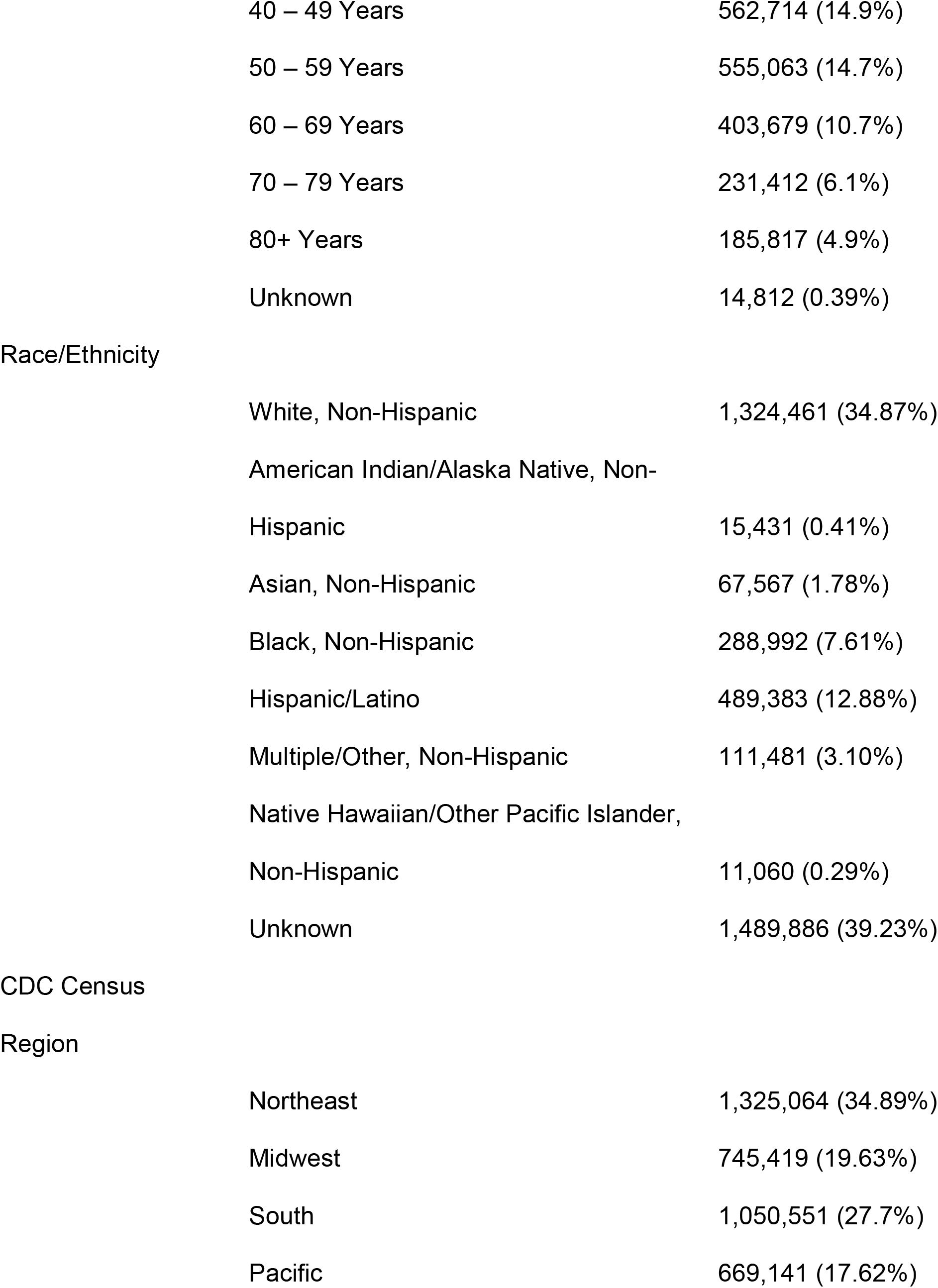
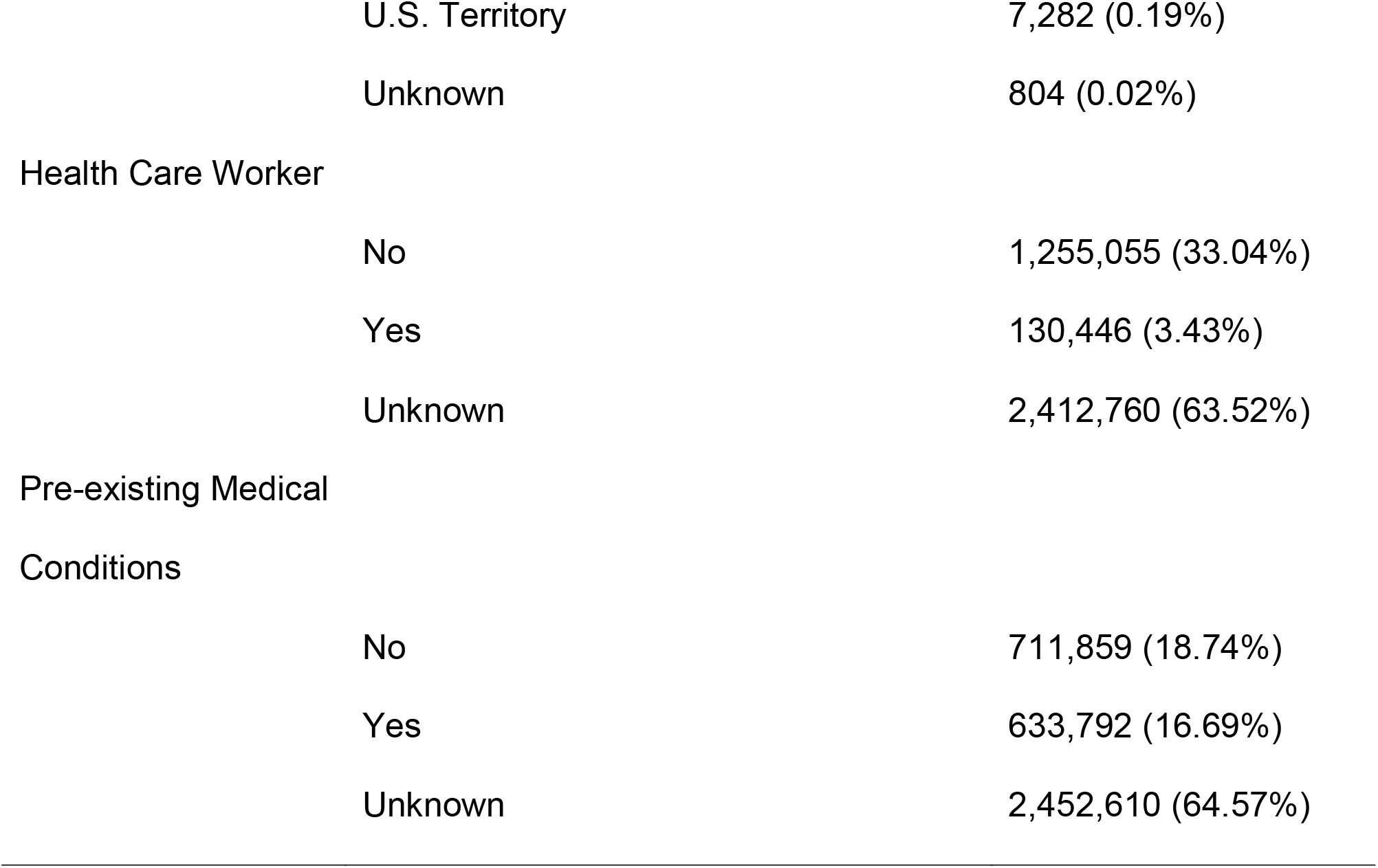
Demographics of patients with COVID-19.

| Variable | Label | Patients with COVID-19<br>(N = 3,798,261) |
| --- | --- | --- |
| Sex |  |  |
|  | Male | 1,795,160 (47.6%) |
|  | Female | 1,954,915 (51.9%) |
|  | Other or Unknown | 18,709 (0.50%) |
| Age Group |  |  |
|  | 0 – 9 Years | 141,591 (3.70%) |
|  | 10 – 19 Years | 379,438 (10.0%) |
|  | 20 – 29 Years | 711,770 (18.8%) |
|  | 30 – 39 Years | 611,965 (16.2%) |

|  |  |
| --- | --- |
| 40 – 49 Years | 562,714 (14.9%) |
| 50 – 59 Years | 555,063 (14.7%) |
| 60 – 69 Years | 403,679 (10.7%) |
| 70 – 79 Years | 231,412 (6.1%) |
| 80+ Years | 185,817 (4.9%) |
| Unknown | 14,812 (0.39%) |

Race/Ethnicity
|  |  |
| --- | --- |
| White, Non-Hispanic | 1,324,461 (34.87%) |
| American Indian/Alaska Native, Non-Hispanic | 15,431 (0.41%) |
| Asian, Non-Hispanic | 67,567 (1.78%) |
| Black, Non-Hispanic | 288,992 (7.61%) |
| Hispanic/Latino | 489,383 (12.88%) |
| Multiple/Other, Non-Hispanic | 111,481 (3.10%) |
| Native Hawaiian/Other Pacific Islander, Non-Hispanic | 11,060 (0.29%) |
| Unknown | 1,489,886 (39.23%) |

Region
|  |  |
| --- | --- |
| Northeast | 1,325,064 (34.89%) |
| Midwest | 745,419 (19.63%) |
| South | 1,050,551 (27.7%) |
| Pacific | 669,141 (17.62%) |
|  | U.S. Territory | 7,282 (0.19%) |
|  | Unknown | 804 (0.02%) |
| Health Care Worker |  |  |
|  | No | 1,255,055 (33.04%) |
|  | Yes | 130,446 (3.43%) |
|  | Unknown | 2,412,760 (63.52%) |
| Pre-existing Medical<br>Conditions |  |  |
|  | No | 711,859 (18.74%) |
|  | Yes | 633,792 (16.69%) |
|  | Unknown | 2,452,610 (64.57%) |

Table 2 presents the six severe COVID-19 outcomes labeled in the CDC dataset. We found that providers hospitalized 13.52% of patients with COVID-19, admitted 4.98% of patients to the ICU, and intubated or mechanically ventilated 3.02% of patients. Additionally, 6.36% of patients with COVID-19 eventually developed pneumonia, 1.78% developed ARDS, and 4.87% eventually died.

**Table 2.** Severe COVID-19 outcomes.

| Outcome | Label | Patients with COVID-19<br>(N = 3,798,261) | Missing Data |
| --- | --- | --- | --- |
| Hospitalization |  |  | 53.5% |
|  | No | 1,526,273 (86.5%) |  |
|  | Yes | 238,638 (13.5%) |  |
| ICU Admission |  |  | 79.3% |
|  | No | 747,280 (95.0%) |  |
|  | Yes | 39,152 (5.0%) |  |
| Mechanical Ventilation/Intubation |  |  | 84.1% |
|  | No | 585,626 (97.0%) |  |
|  | Yes | 18,244 (3.0%) |  |
| Pneumonia |  |  | 79.1% |
|  | No | 741,769 (93.6%) |  |
|  | Yes | 50,373 (6.4%) |  |
| Acute Respiratory Distress |  |  | 79.2% |
|  | No | 777,033 (98.2%) |  |
|  | Yes | 14,054 (1.8%) |  |
| Death |  |  | 54.6% |
|  | No | 1,638,857 (95.1%) |  |
|  | Yes | 83,810 (4.9%) |  |

To understand the risks of severe COVID-19 outcomes in special populations, we performed a univariate subgroup analysis on healthcare providers and subjects with pre-existing conditions. Table 3 demonstrates that patients with pre-existing medical conditions had a higher risk of developing all six severe COVID-19 outcomes than the opposing group. Likewise, this table also illustrates that healthcare workers have a significantly decreased risk of obtaining all six severe COVID-19 outcomes than non-healthcare workers. To determine the risk of severe outcomes by age, we plotted the age groups by the proportion of patients with severe COVID-19 in Fig 1. This figure demonstrates that all six outcomes increase in older subjects, while death and hospitalizations escalate at a steeper rate.

**Table 3.** Severe COVID-19 outcomes in special populations.

| Outcomes | Medical Conditions<br>(N = 633,792) | Healthcare<br>Workers<br>(N = 130,446) | Overall<br>(N = 3,798,261) |
| --- | --- | --- | --- |
| Hospitalized | 125,449 (23.9%) * | 4,191 (4.2%) * | 238,638 (13.5%) |
| ICU Admission | 25,656 (8.9%) * | 774 (1.6%) * | 39,152 (5.0%) |
| Mechanical Ventilation | 12,964 (5.3%) * | 301 (0.7%) * | 18,244 (3.0%) |
| Pneumonia | 34,935 (10.9%) * | 2,020 (4.1%) * | 50,373 (6.4%) |
| Acute Respiratory Distress | 10,397 (3.3%) * | 485 (1.0%) * | 14,054 (1.8%) |
| Death | 46,449 (10.3%) * | 233 (0.2%) * | 83,810 (4.9%) |

\* Proportions are different compared to overall population (p < 0.001)

**Fig 1.**
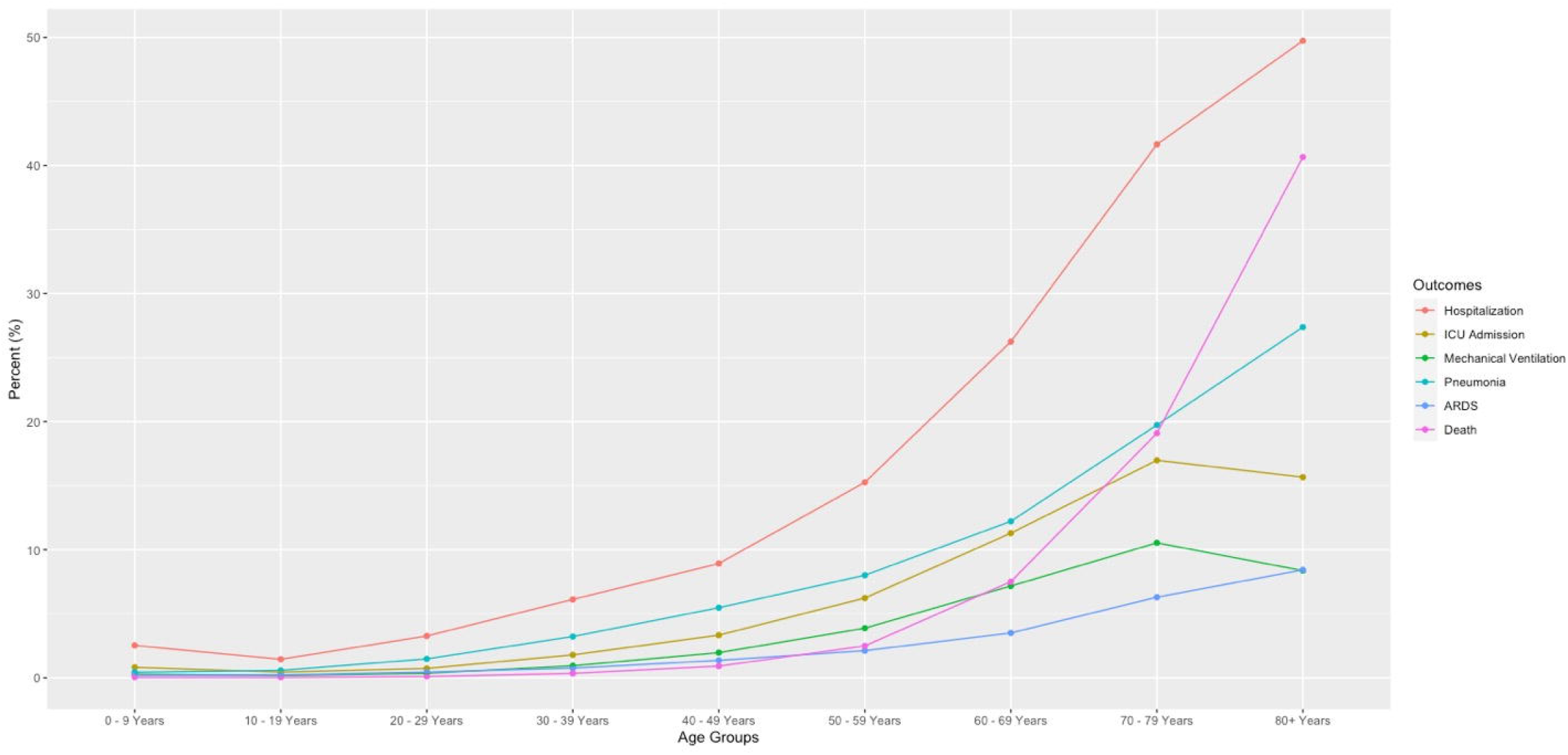
Risk of severe COVID-19 by age group.

We accounted for potential confounding factors in a multivariate logistic regression model for each of the six severe outcomes controlling for the same seven independent variables of sex, age group, race/ethnicity, healthcare worker status, U.S. Census region, pre-existing medical condition status, and month of positive COVID-19 test (Figure 2). We compared the logistic regression model using all variables to the LASSO selected models and presented the best model metrics to evaluate predictive capacity.

**Fig 2.**
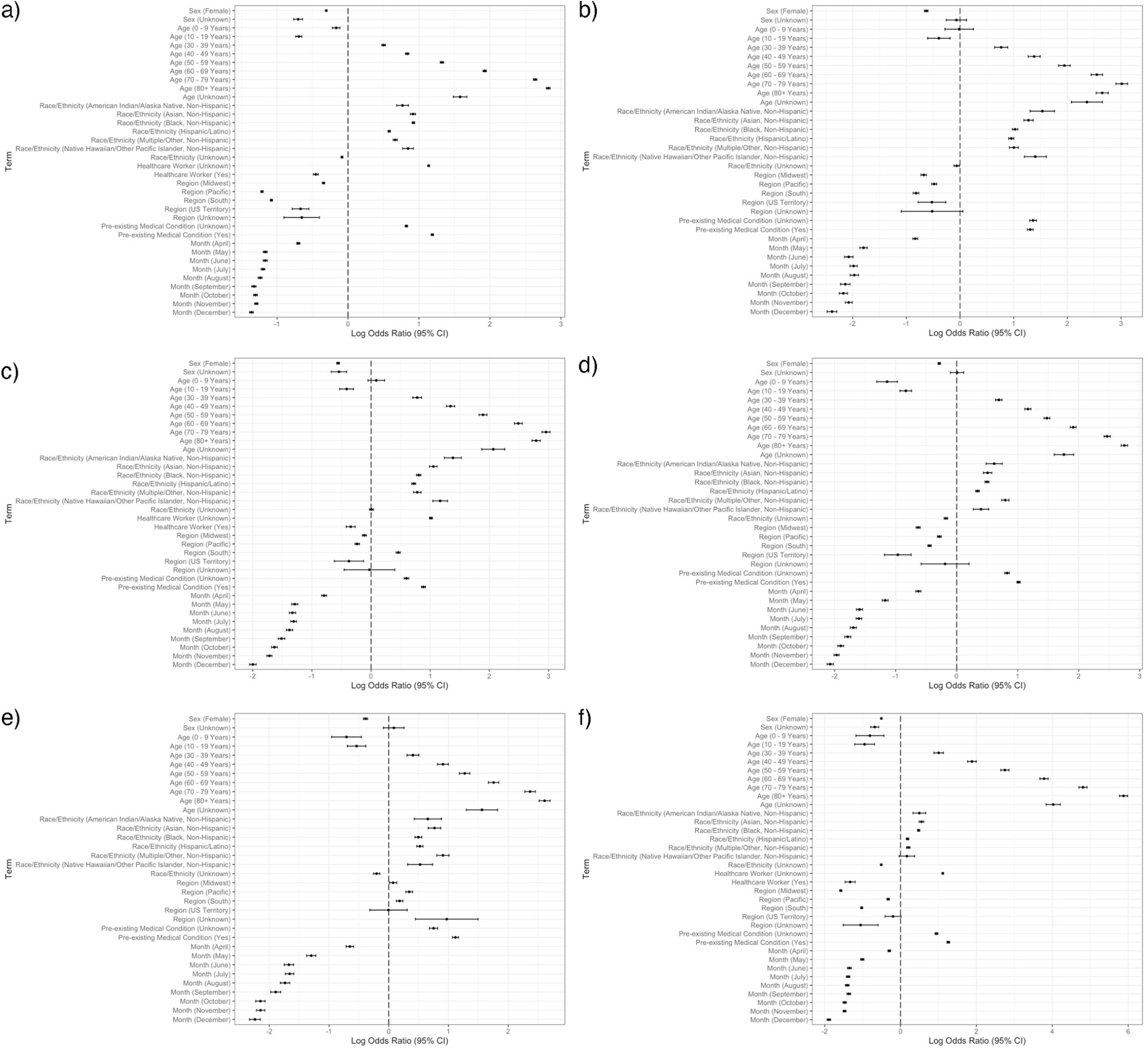
Logistic regression models for severe COVID-19 outcomes. Forest plot for each severe COVID-19 outcome. a) Hospitalization – all variables, b) ICU admission – all variables, c) Mechanical ventilation/intubation – LASSO selected variables, d) Pneumonia – LASSO selected variables, e) Acute respiratory distress – LASSO selected variables, and f) Mortality – all variables. Male is the reference group for sex. Age 20-29 Years is the reference group for age. Non-Hispanic White is the reference group for race/ethnicity. Northeast is the reference group for the region. The reference group for the month of the positive test is January-March.

Figure 2a illustrates the results of the logistic regression model created for the hospitalization outcome. The LASSO algorithm did not drop any of the variables, and therefore, we did not create its associated model (S1 Table). When controlling for all other covariates, we found the three largest effect sizes were in patients over 80, between the ages of 70-79, and with pre-existing medical conditions. The forest plot shows that patients over 80 and 70-79 years old had seventeen- and fourteen-times higher odds of being hospitalized than those with ages 20-29, respectively. Patients with a pre-existing medical condition had a three-fold increased risk of being hospitalized.

Additionally, the LASSO algorithm did not drop any variables while creating the ICU admission logistic regression model (Fig 2b, S2 Table). Patients ≥ 80 and between the ages of 70-79 had a seventeen- and twenty-times increased odds of admission to the ICU, respectively. American Indian/Alaska Native, non-Hispanic patients had a four times greater risk of ICU admission likelihood than white, non-Hispanic patients. Patients with a pre-existing medical condition had a four times greater risk of ICU admission.

In the outcome of mechanical ventilation or intubation, the LASSO algorithm dropped the variable healthcare worker status (Fig 2c). However, compared to the logistic regression model with all variables, the model fit metrics were nearly equivalent, suggesting that the LASSO model did not significantly improve the model fit for mechanical ventilation (S1 Fig, S3 Table). Fig 2c illustrates the forest plot for the LASSO selected variables for the mechanical ventilation logistic regression models. The characteristics with the most significant effect sizes include patients 70-79 years old, 60-69 years old, and those with pre-existing medical conditions. Patients in their seventh and eighth decades of life had a 17- and 12-fold increase risk of being intubated or put on mechanical ventilation, respectively. Subjects with pre-existing medical conditions had a threefold increase of being intubated or ventilated.

The LASSO model for pneumonia also dropped healthcare workers as a variable to include; however, compared to the all-variables model, the metrics were relatively equivalent, suggesting that dropping the variable did not improve model fit (S2 Fig S4 Table). Fig 2d presents the forest plot for the LASSO selected logistic regression model. Patients over the age of 80 and between the ages of 70-79 had the most significant effect sizes with an increased odds of developing pneumonia by 17- and 12-fold, respectively. Patients with pre-existing medical conditions had the third-highest effect size with an OR of 2.8.

The LASSO algorithm for ARDS also dropped healthcare workers from its model; however, compared to the logistic regression with all variables, the variables’ effect sizes and p-values were similar (S3 Fig, S5 Table). The forest plot in Fig 2e presents the ORs of the logistic regression for the LASSO selected variables. Patients over 80 and in their seventh decade of life had a fifteen- and eleven-times increased risk of developing ARDS compared to the 20–29-year age group, respectively. After age groups, the third-largest effect size demonstrates that patients with pre-existing medical conditions have increased ARDS odds by three.

The LASSO model for mortality did not drop any variables (S6 Table). We present the mortality regression models with all variables in Fig 2f. Patients older than 80 years old and between the ages of 70-79 had the two largest effect sizes. Patients over 80 and in the seventh decade of life have 356- and 122-times increased risk of mortality, respectively, compared to patients aged 20-29. Pre-existing medical conditions had the third-highest effect size with a fourfold increased risk of mortality.

Next, we created a prediction model using the same independent variables to calculate a patient’s risk of severe COVID-19 outcomes if they were to contract the disease. We present the hyperparameter tuning results from 3-fold cross-validation on the training sets in S7 Table. For each outcome, an initial grid search across a broader range of values led to an optimum number of variables available for splitting at each tree node, or mtry value of 1 and a minimum number of data points required for the node to be split further, or min_n value of 18 leading to the best performance. The grid search was then repeated on a narrower range of hyperparameter values to produce the ROC curves for each outcome.

The ROC curves for the final random forest models fit the entire training set using the optimal hyperparameter values for each outcome shown in Figure 3. All of the models had a high AUC > 0.8. The model with death as the outcome had the highest performance (AUC = 0.953), followed by hospitalization (AUC = 0.892) and mechanical ventilation (AUC = 0.882).

**Fig 3.**
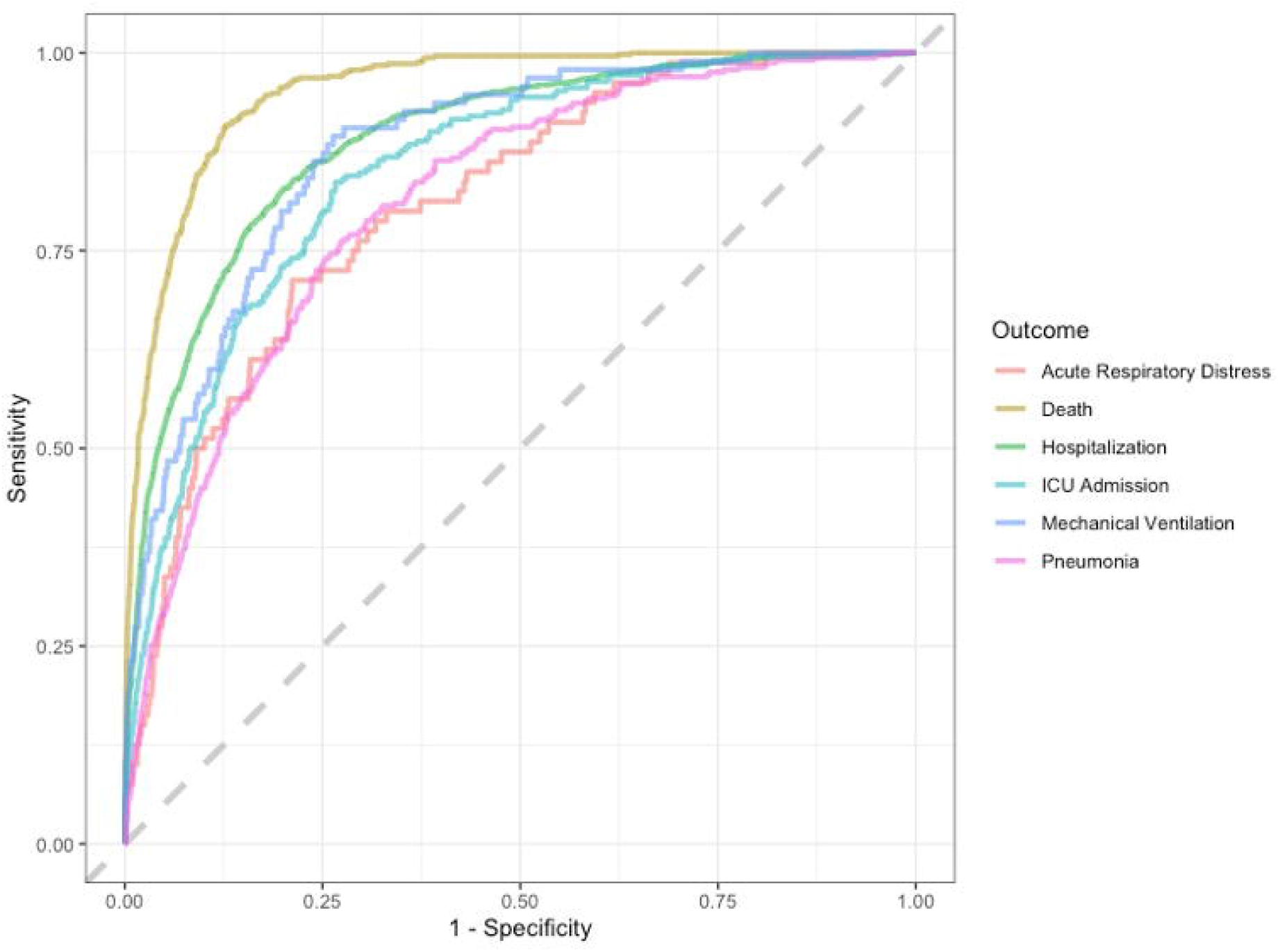
ROC curves of random forest models. Abbreviation = ROC, receiver operating characteristic curve

Table 4 presents the performance metrics for both the random forest and LASSO models for each outcome. Relative to the LASSO models, the random forest models had a higher performance in overall accuracy for each outcome. The models for predicting mechanical ventilation and deaths had higher accuracy than predicting pneumonia and ARDS. The random forest models for hospitalizations and death had a strong recall, with a low number of false-negative predictions. Overall, we found that the random forest models had a low precision with a high number of false positives. We presented the relative feature importance for each model in Fig 4 to rank the patient characteristics that had the most influence on each outcome. Age groups and presence of a pre-existing condition were the top features for each outcome. Northeast region was an important feature in predicting hospitalization or ventilation. This pattern draws upon high hospitalization and ventilation rates in the Northeast during the first COVID-19 wave in the spring. The youngest age group is likely a solid indicator for the model of a lack of the outcome occurring.

**Table 4.**
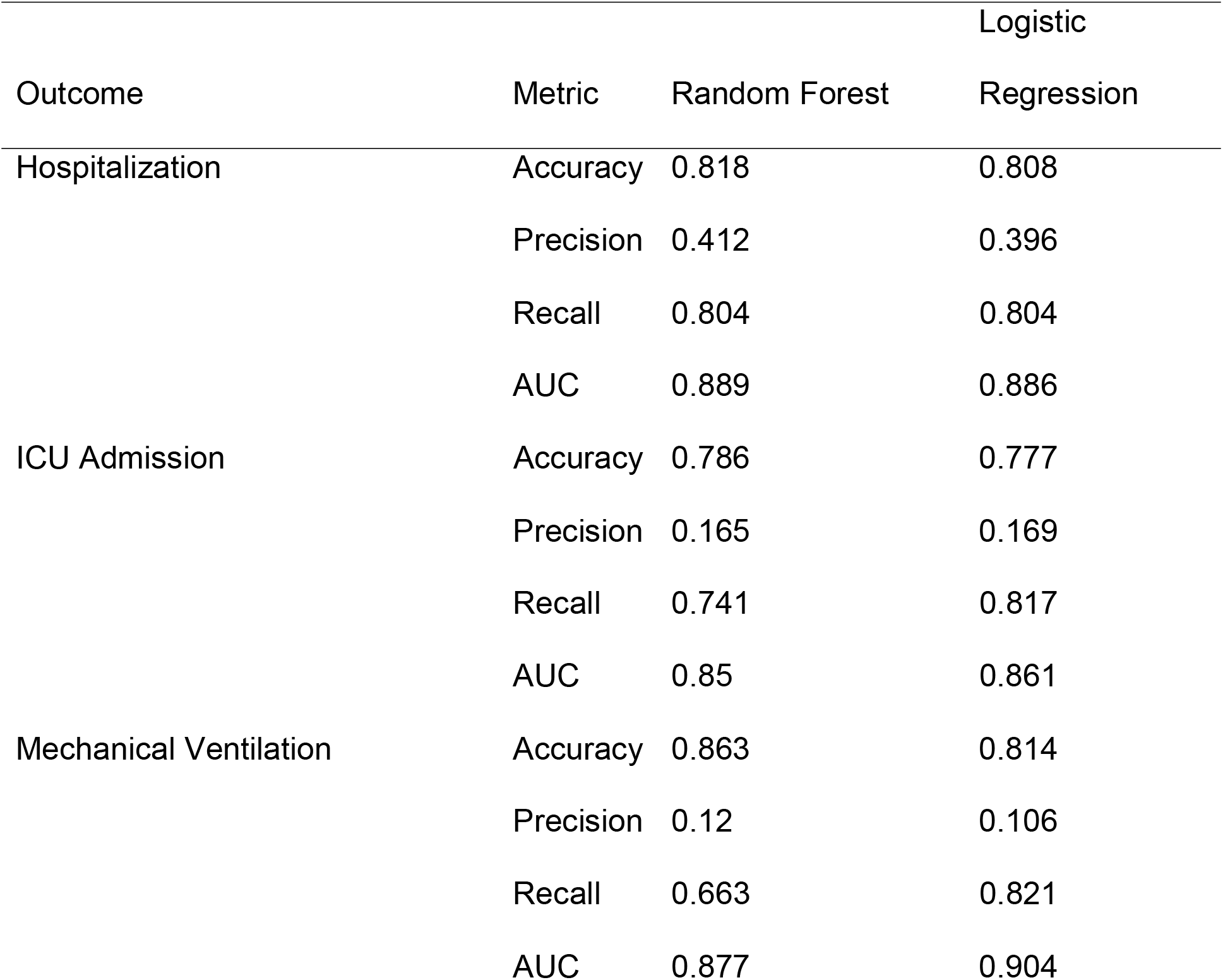
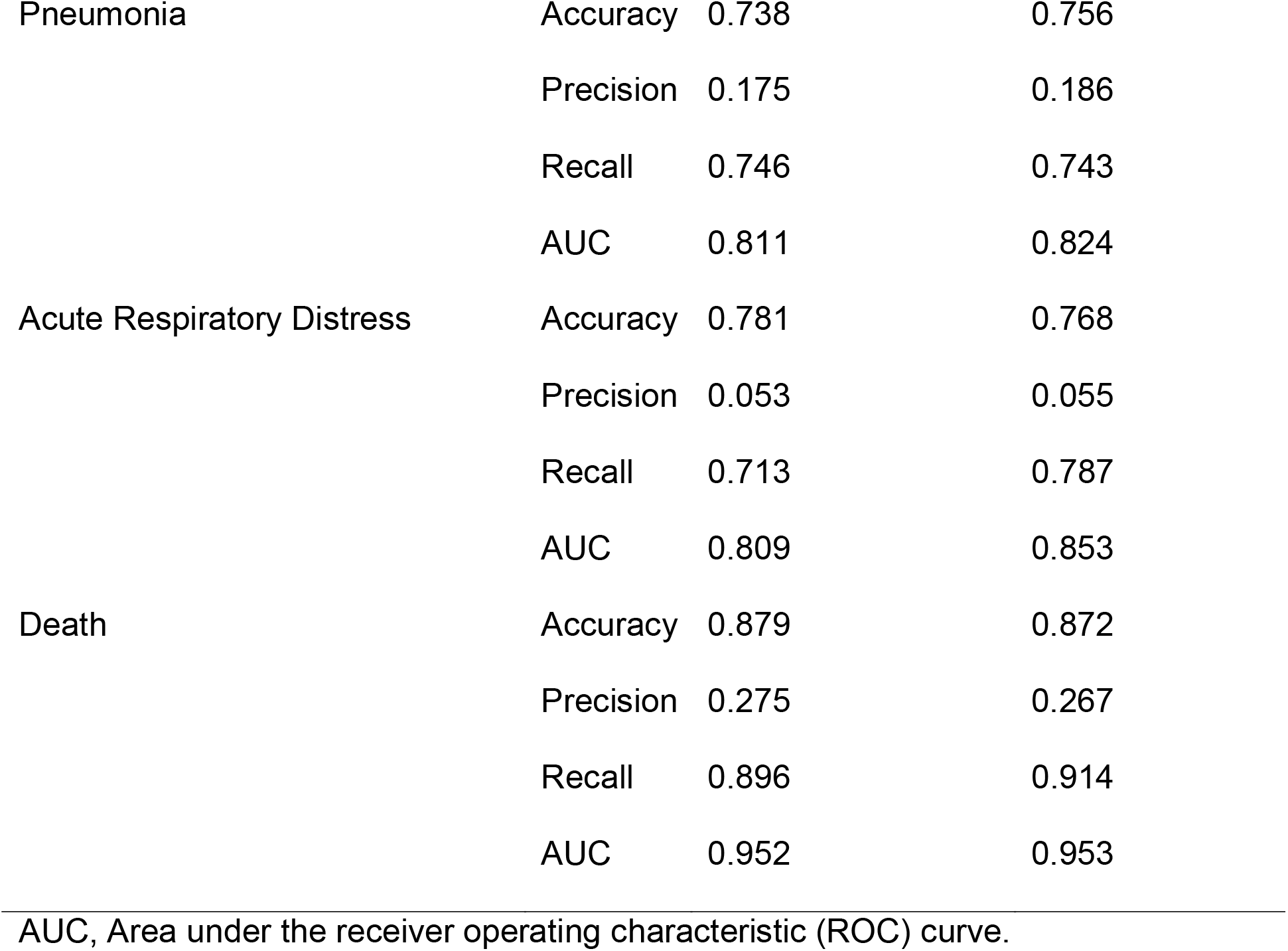
Performance metrics.

**Fig 4.**
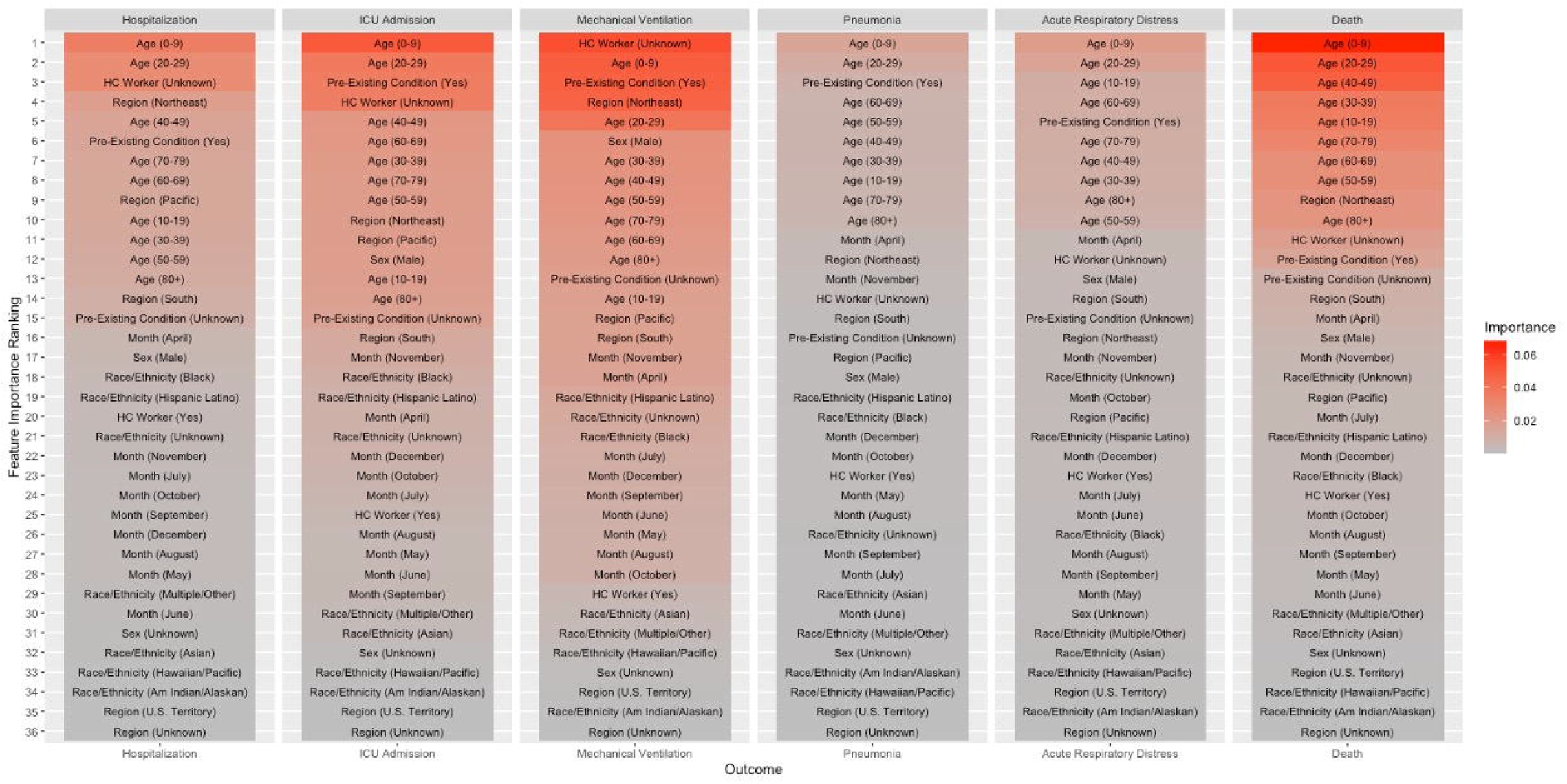
Figure importance by the outcome. Abbreviation = HC Worker, Healthcare Worker.

We aggregated the predictive models into the Severe COVID-19 Calculator web application and published it online (https://methodsconsultants-apps.shinyapps.io/guthrie-cdc-covid-prediction/). The web application accounts for all predictive variables and provides a predicted estimate of the probability of severe COVID-19 outcomes (Fig 5).

**Fig 5.**
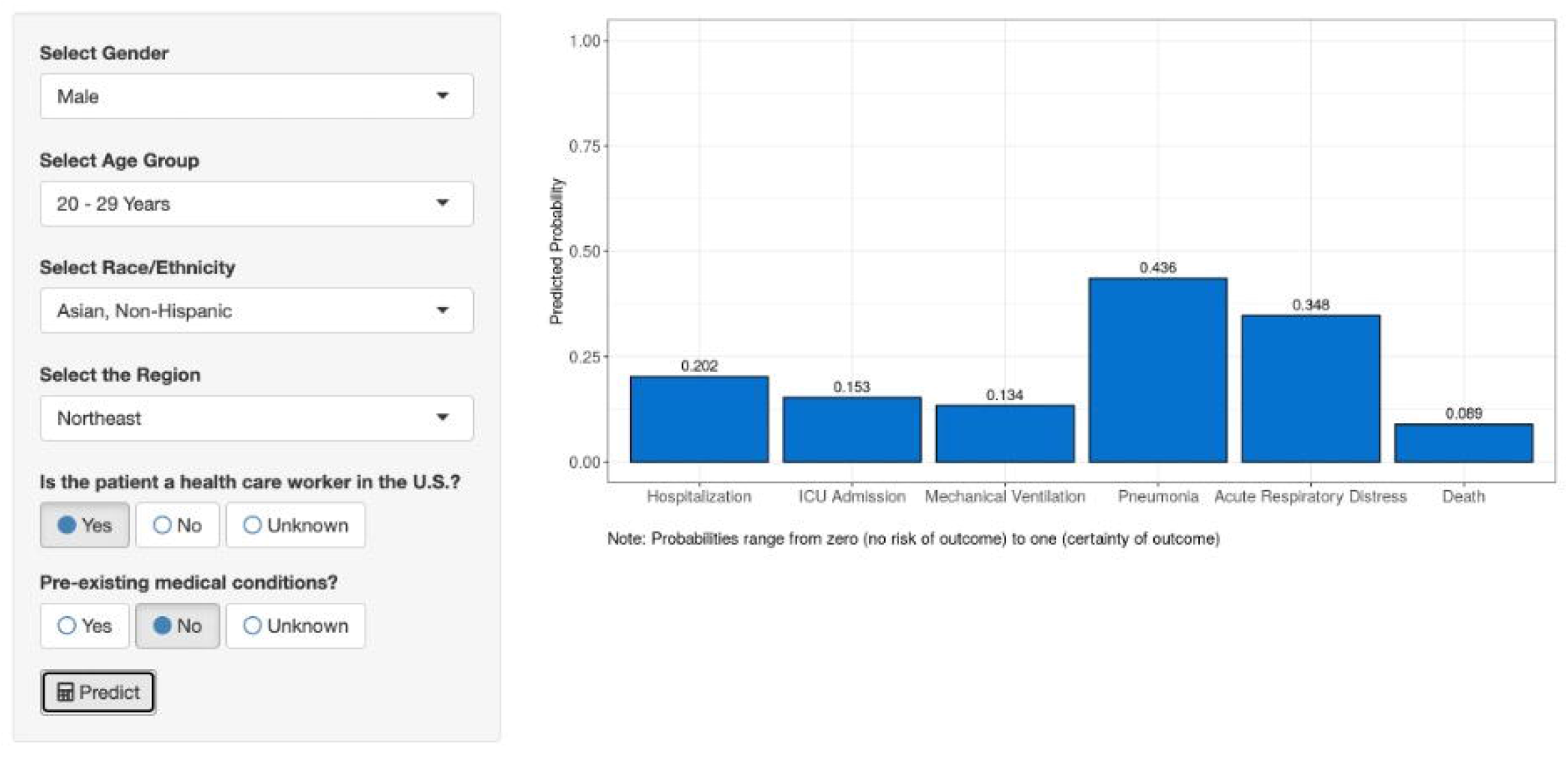
Example of the Severe COVID-19 Calculator.

## Discussion

Among 3,798,261 patients with COVID-19 from the pre-vaccine era in the United States, older patients had an increased risk of all six severe outcomes. The odds ratios are significantly higher in the latter decades of life as some outcomes, such as death, have a 300-fold increase. We discovered that White subjects had a decreased occurrence of all severe outcomes than Non-Whites, except Native Hawaiian/Other Pacific Islander patients who had similar mortality risks. The models also demonstrated that the rates of severe COVID-19 effects were unequal between all five of the CDC Census Regions. In both the LASSO selected and all-variable models, we found that healthcare workers had a decreased risk of severe COVID-19 outcomes. Additionally, we discovered the patients with pre-existing health conditions all had an increased chance of severe outcomes. When we adjusted for the diagnosis time, we uncovered that severe COVID-19 outcomes decreased by month as the pandemic progressed.

Overall, our results confirm the current COVID-19 literature on severe outcomes. We found that age has the most significant effect measure compared to our other covariables. However, instead of using threshold ages, our age groups provide more details on risk stratification in special populations such as school children. A correspondence published in the New England Journal of Medicine examining child and teacher morbidity in Sweden without school closure also confirmed the low incidence of severe COVID-19 among schoolchildren and preschool-age children during the pandemic [21]. While most researchers confirmed the effects of chronic diseases on severe COVID-19, our results demonstrate that pre-existing conditions have the second most significant influence on outcomes after age [22]. These chronic conditions like obesity and diabetes demonstrate an increased clinical severity in disease presentations, while infrastructure shutdown delayed treatments in conditions such as mental health disorders, chronic kidney disease, and cancer [22]. Additionally, our results also demonstrate that race/ethnicity has a significant influence on severe outcomes. A report from PNAS presented a disproportionate mortality rate in the Black and Latino populations compared to White patients [16]. However, we discovered that all Non-White populations, in general, had higher rates of severe outcomes. Additionally, we found that while healthcare workers consisted of 3% of our sample, they had a decreased risk of severe COVID-19 outcomes or had no influence on their outcomes in this pre-vaccine era. Our results differ from the current idea that healthcare workers mirror the general population rates of severe outcomes [23]. Because a few models excluded the variable healthcare workers, we suspect that the decreased occurrence of severe outcomes reflects a younger and healthier subgroup compared to the general population. A systematic review of COVID-19 in healthcare workers worldwide mirrors our results as providers over 70 or males had a higher risk of death [23].

We present the first report on the CDC COVID-19 case surveillance system. This restricted database presents individual-level data from autonomous reporting entities from all U.S. states and territories. The large sample size and standardized data dictionary allowed us to perform multiple logistic regression models to control each predictive variable. Therefore, we are confident in our adjusted risk calculations as they already account for any potential interactions. To the best of our knowledge, this is the first study to stratify by date of COVID-19 diagnosis to account for any treatment biases as COVID-19 management has changed over time. Additionally, as states have different COVID-19 protocols and strains occupy different regions of the U.S., our study is the first to account for these factors by accounting for geographical regions.

Our web app for our Severe COVID-19 Calculator allows anyone to estimate the risk of the severe outcome if they have or if they were to contract COVID-19. As mentioned throughout our report, all of the predictive variables discussed significantly influence severe COVID-19 outcomes. Thereby, our calculator can provide accurate numeric predictions using the subject’s objective data without arbitrary thresholds or scoring systems. For example, Fig 5 illustrates the user interface and outcome estimates for a hypothetical patient.

While our study accounted for socioeconomic factors such as sex, race, and region, the available data limited our ability to account for other important social determinants of health, such as income and occupation, which may affect severe COVID-19 outcomes and vaccine distribution plans [24]. Additionally, to preserve patient anonymity, we could not distinguish between types of pre-existing conditions. This limitation also questions the possibility of the number of pre-existing conditions correlating with disease severity. If so, there may be a benefit in making this distinction in future predictive tools.

Like all prediction modeling projects, the exclusion of potentially essential variables could reduce our models’ performance. Also, within the available data, a few predictive variables, such as race/ethnicity, healthcare worker status, pre-existing medical conditions, contained unknown variables, which could have affected our calculator’s accuracy. However, the high AUC value and performance metrics for all of our outcomes suggest that our predictive models are significantly better than random chance.

As vaccine distribution continues to roll out in phases and trends of severe COVID-19 outcomes change, we will need to continuously recalculate our results to determine the patients at the highest risk for severe COVID-19 outcomes. With the continuously updated CDC data and our publicly available web app, we can recreate our Severe COVID-19 Calculator to reflect the most up-to-date data. For example, in a new data release, we may find a race/ethnicity, sex, or age group with a disproportionate chance of severe COVID-19 outcomes in the vaccination era, and we would like to account for that in future calculator iterations. Additionally, using some of the variables already available in the CDC report, we plan to aggregate data sources to account for variables of interest, such as estimated patient income.

Since the start of the pandemic, researchers have published dozens of studies that identified risk factors for severe outcomes. However, the literature lacked any detail on each risk factor’s relative importance or adjusted for potential covariable interactions. Our study performed these adjustments and found that predicting severe COVID-19 outcomes is a multifactorial problem and quantified that variables such as age, pre-existing health conditions, and race/ethnicity are more important than others like health care worker status.

While we provide an objective tool to assess a subject’s risk of developing severe COVID-19 outcomes, we have no comment on current vaccine distribution plans. The Severe COVID-19 Calculator is an objective tool that mirrors other models already used in medicine to help providers and researchers stratify patients in resource scarcity, such as hospital beds, ventilators, masks, or vaccines. This study’s sole purpose was to identify independent risk factors and quantify these effects to understand the apparent outcome disparities between different patient groups. After adjusting for covariates, patients that are older, male, Non-White, non-healthcare workers, or possess at least one pre-existing condition have an increased risk of severe COVID-19 outcomes. Additionally, we found that our Severe COVID-19 Calculator accurately predicts the chance of hospitalization, ICU hospitalization, mechanical ventilation/intubation, pneumonia, ARDS, and death in citizens that may contract or have COVID-19.

## Supporting information

Letter of Ethical Approval

## Data Availability

No - some restrictions will apply. The data used to generate this manuscript can be found from the CDC Website. As mentioned in our methods, the data is available as "COVID-19 Case Surveillance Restricted Access."

https://data.cdc.gov/Case-Surveillance/COVID-19-CaseSurveillance-Restricted-Access-Detai/mbd7-r32t

## Acknowledgments

We thank Vicky Hickey, Dr. Sri Harshavardhan Senapathi MD, Dr. Seungwoo Chai MD, and Dr. Karen Williams, PharmD., BCPS-AQ ID (Guthrie Robert Packer Hospital) for helping us develop our study design and comments on the manuscript.

## Supporting information

**S1 Fig.**
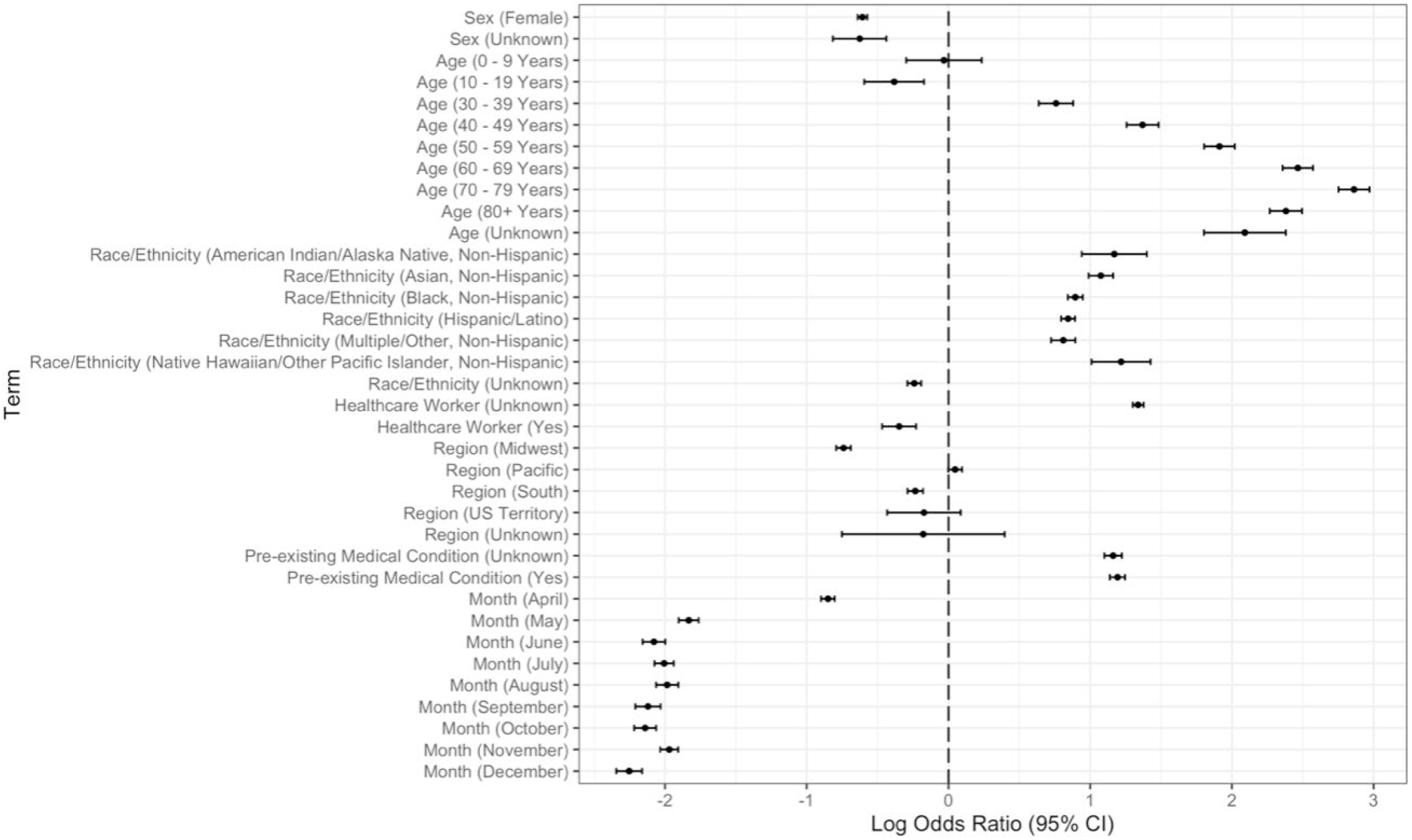
Forest plot for mechanical ventilation logistic regression model with all variables. Male is the reference group for sex. Age 20-29 Years is the reference group for age. Non-Hispanic White is the reference group for race/ethnicity. Northeast is the reference group for the region. The reference group for the month of the positive test is Jan-March.

**S2 Fig.**
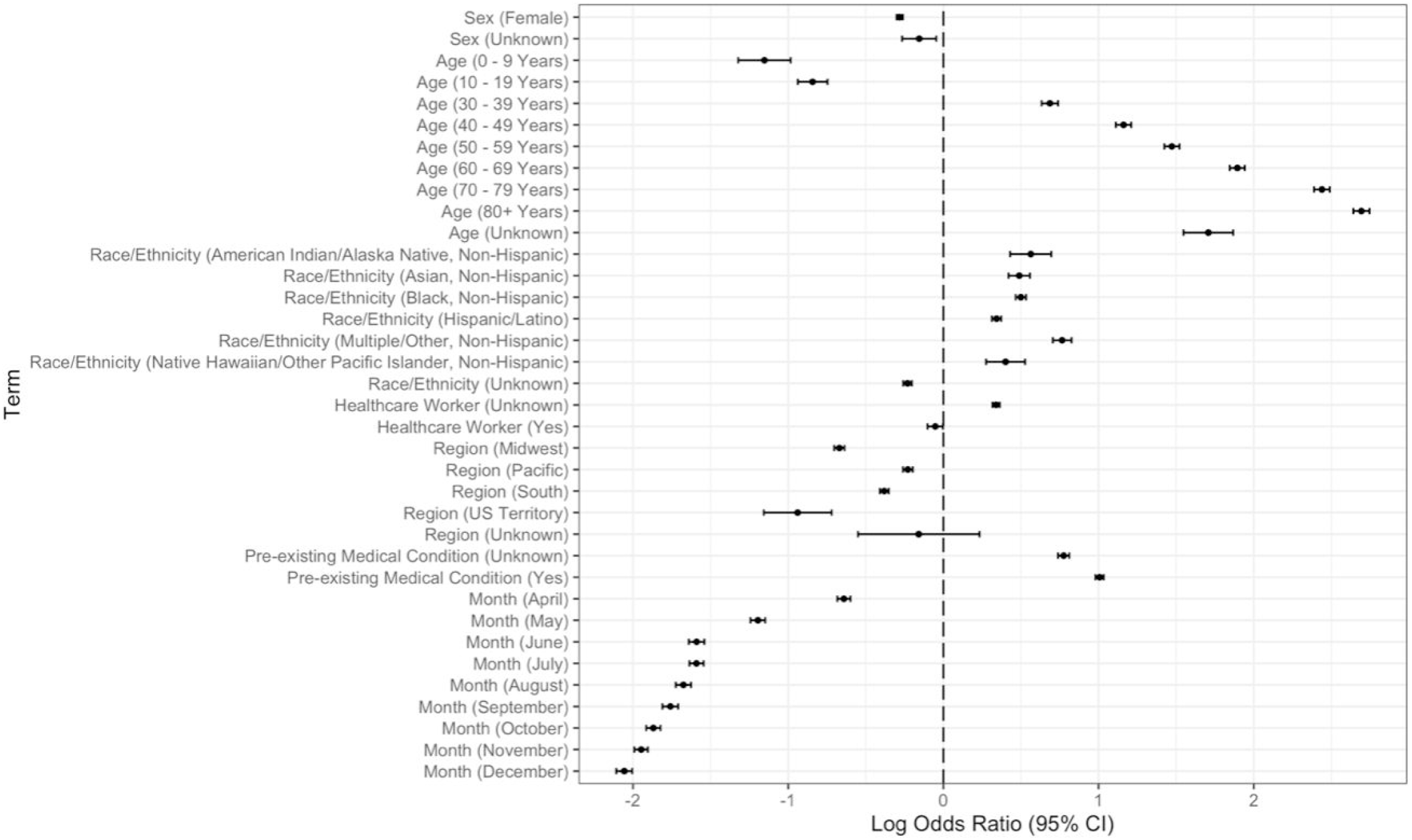
Forest plot for pneumonia logistic regression model with all variables. Male is the reference group for sex. Age 20-29 Years is the reference group for age. Non-Hispanic White is the reference group for race/ethnicity. Northeast is the reference group for the region. The reference group for the month of the positive test is Jan-March.

**S3 Fig.**
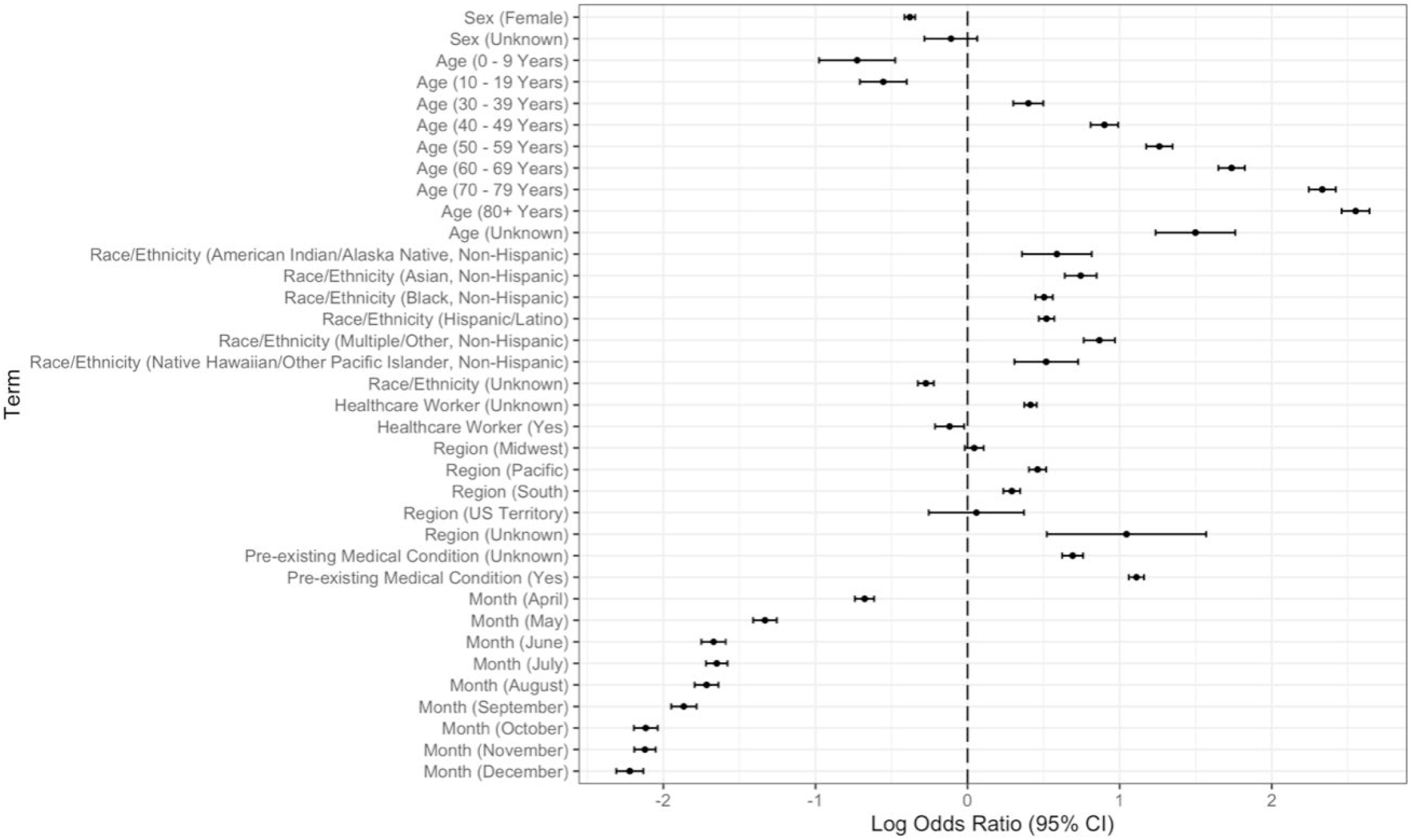
Forest plot for acute respiratory distress logistic regression model – all variables. Male is the reference group for sex. Age 20-29 Years is the reference group for age. Non-Hispanic White is the reference group for race/ethnicity. Northeast is the reference group for the region. The reference group for the month of the positive test is Jan-March.

**S1 Table.**
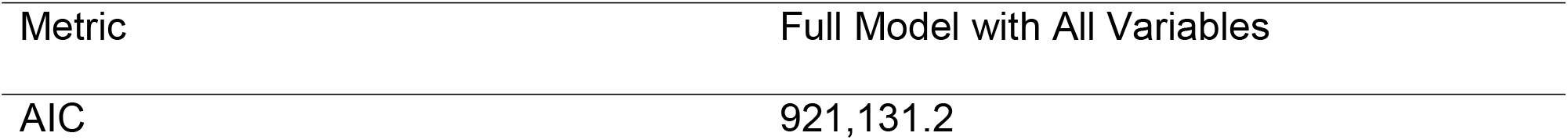
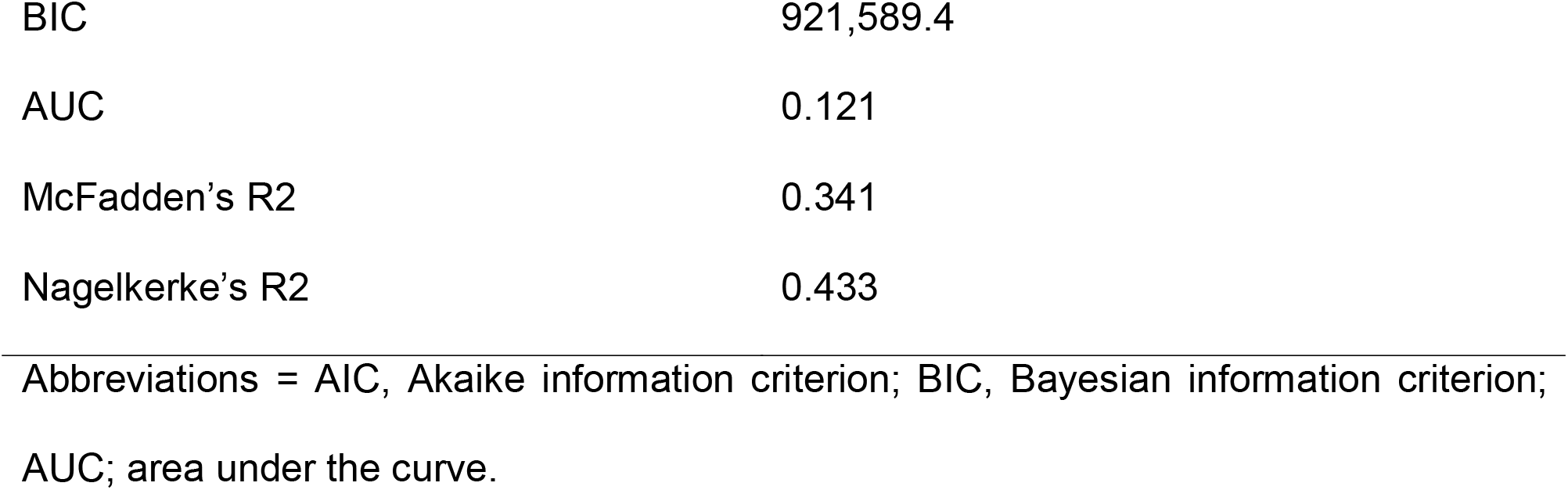
Model fit statistics for hospitalization logistic regression model.

**S2 Table.**
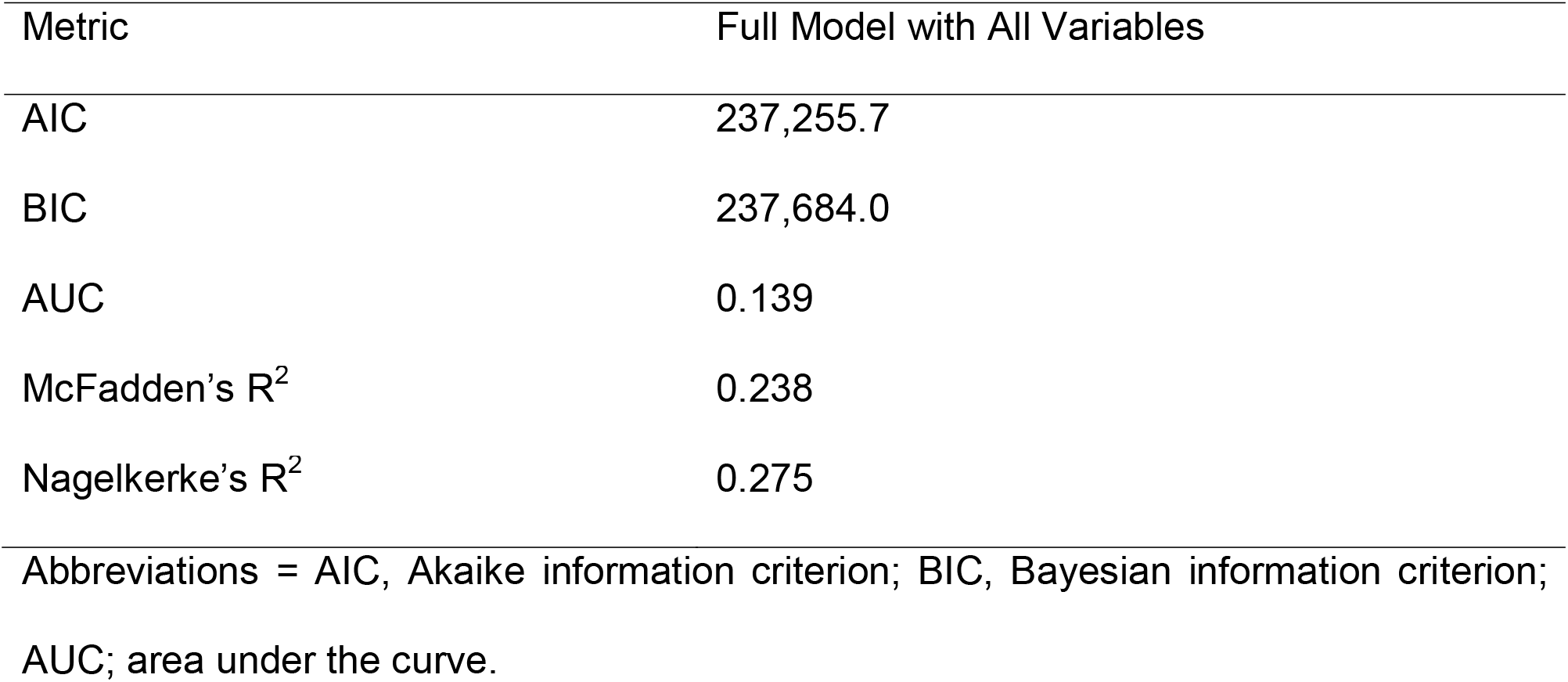
Model fit statistics for ICU admission logistic regression models.

**S3 Table.**
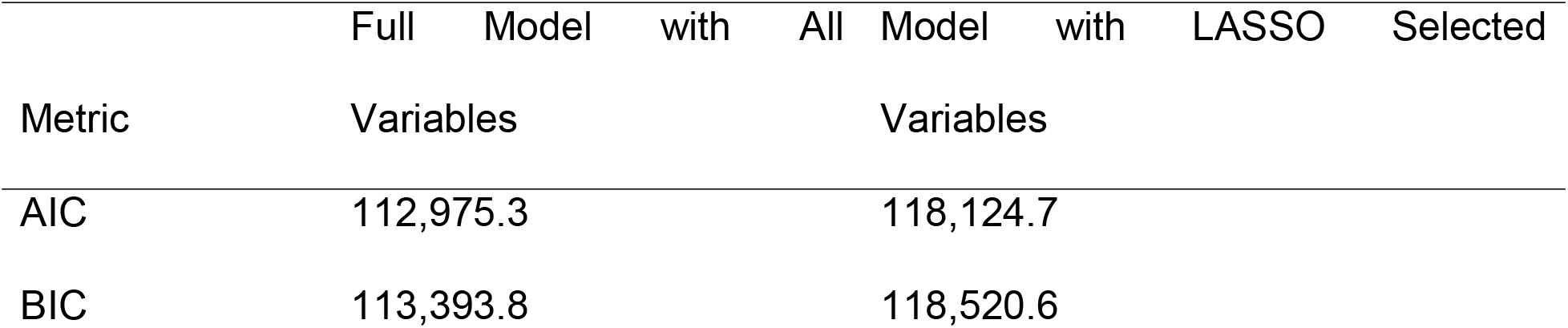
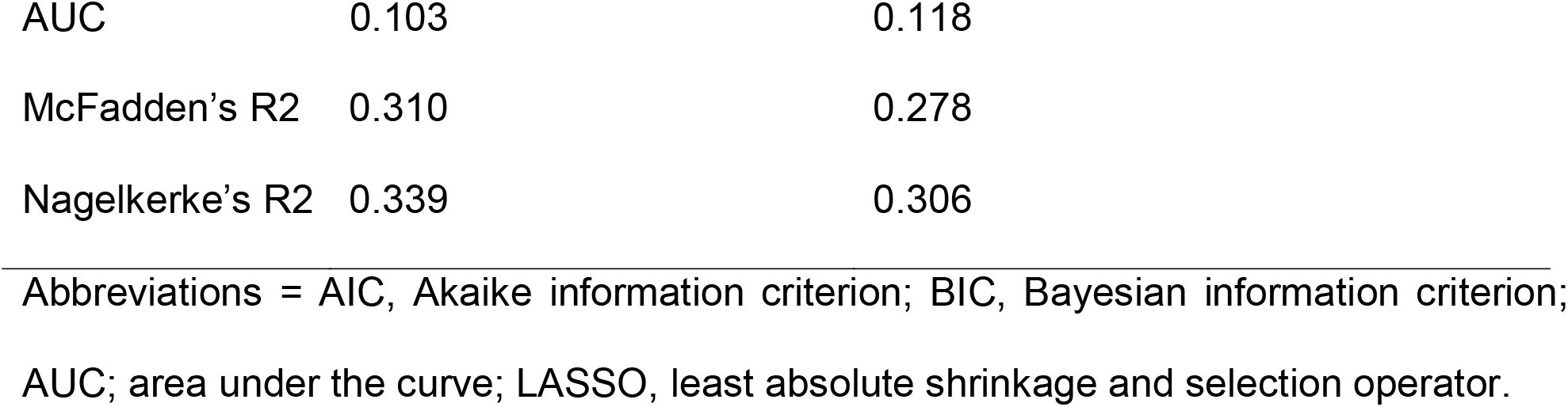
Model fit statistics for mechanical ventilation logistic regression models.

**S4 Table.**
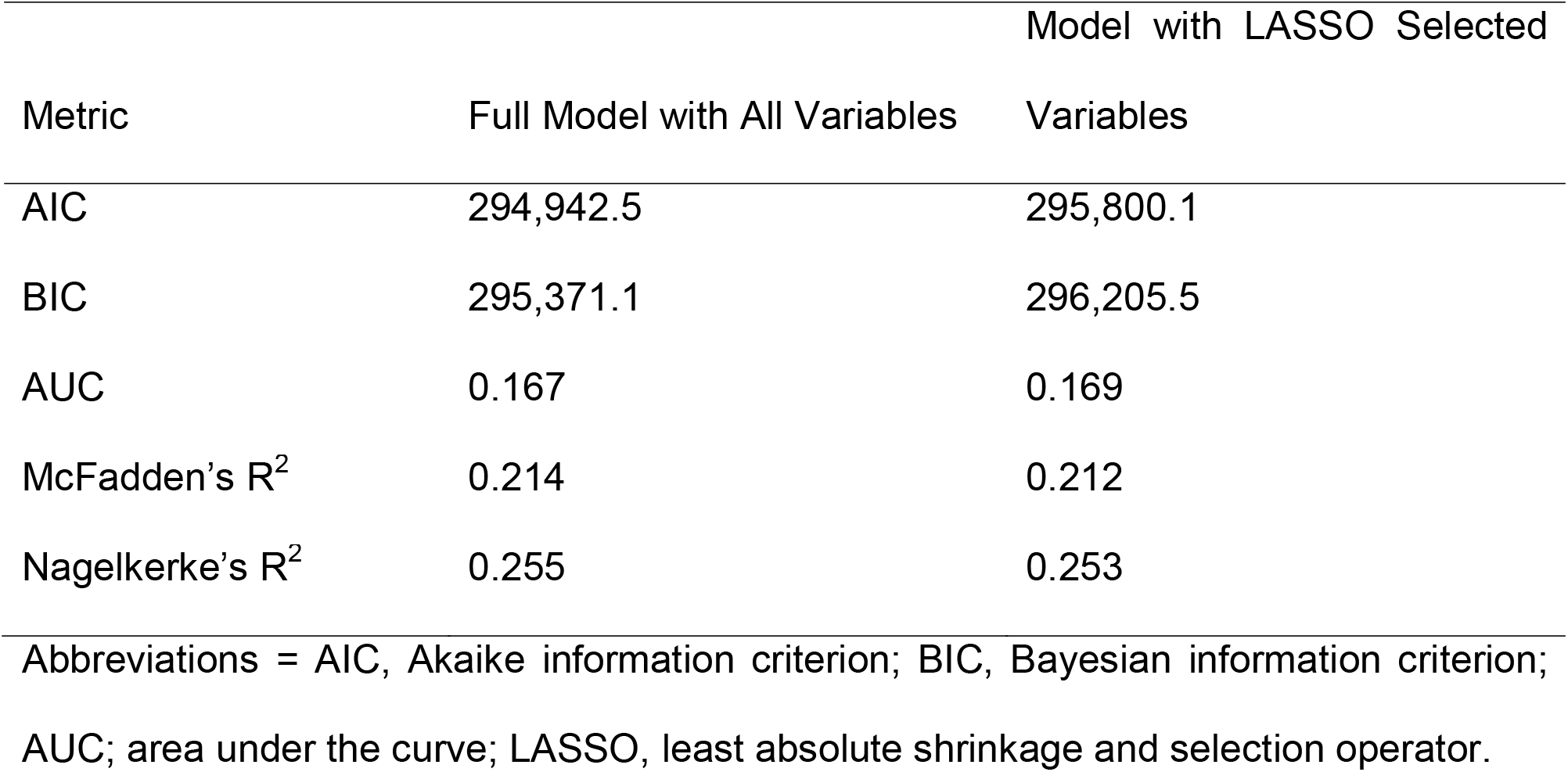
Model fit statistics for pneumonia logistic regression models.

**S5 Table.**
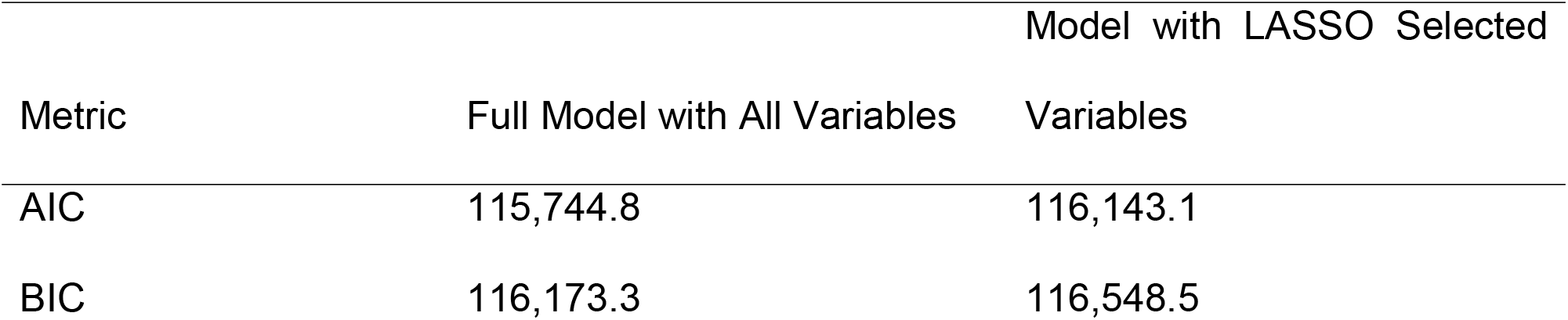
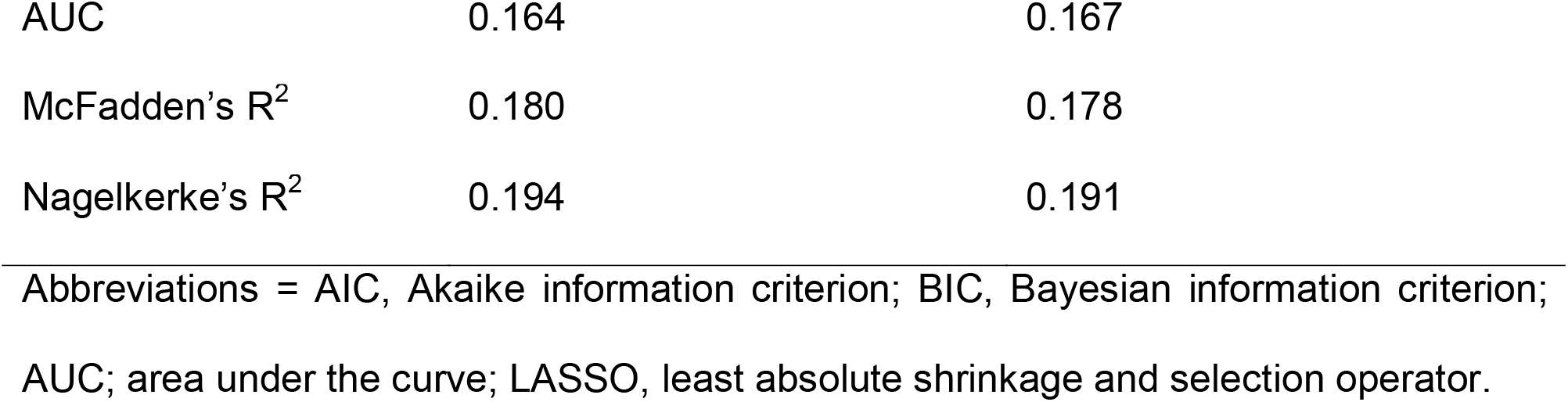
Model fit statistics for acute respiratory distress logistic regression models.

**S6 Table.**
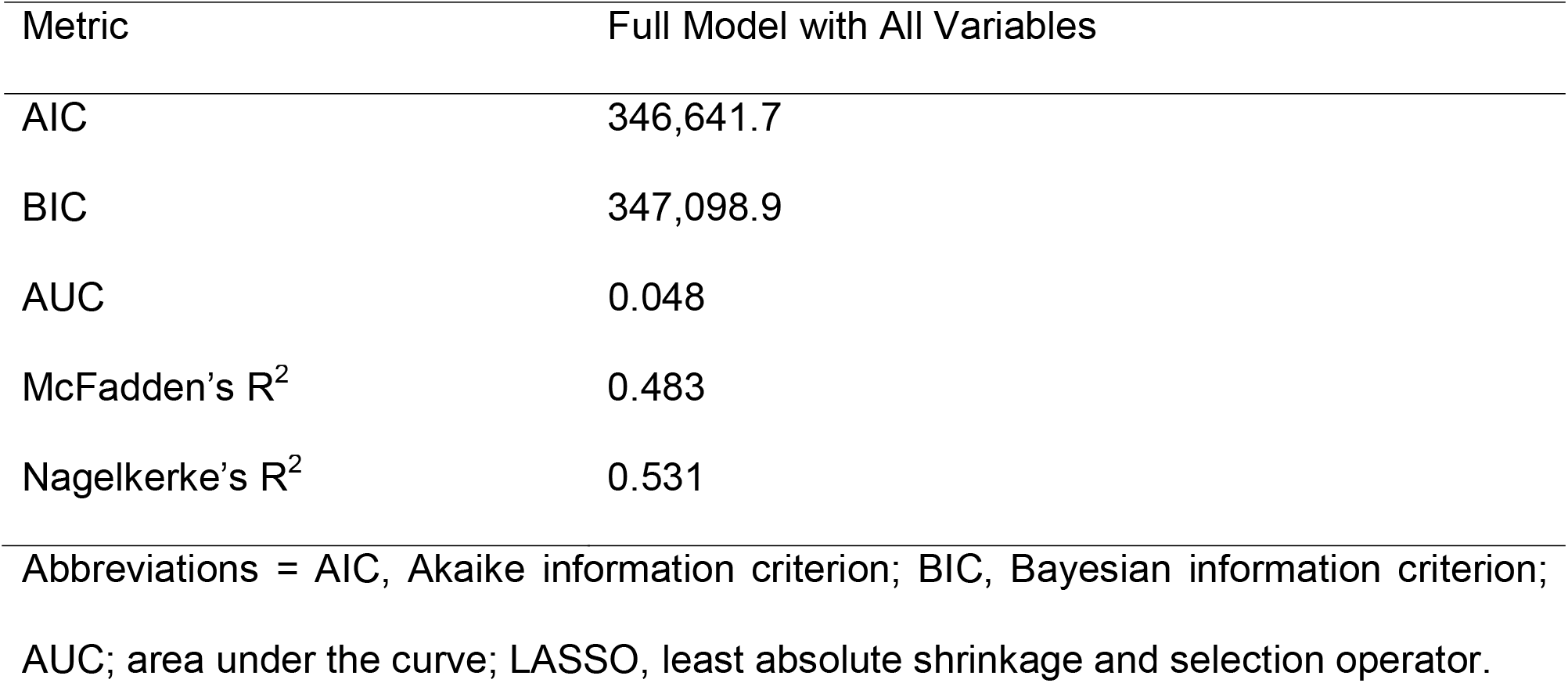
Model fit statistics for mortality logistic regression model.

**S7 Table.**
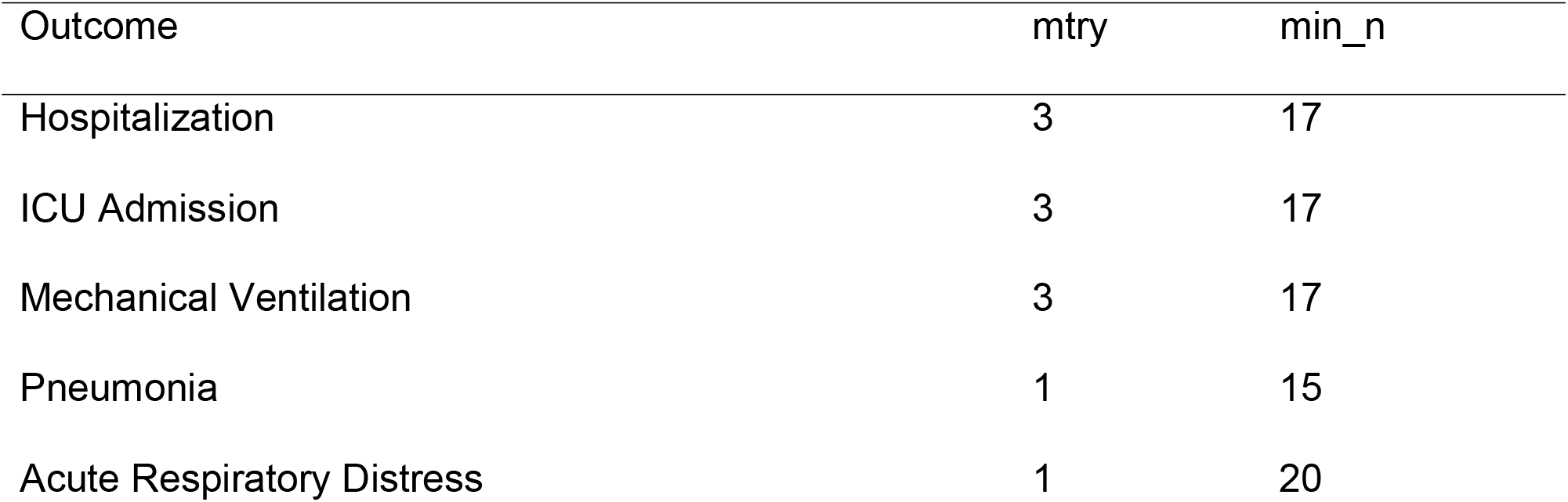
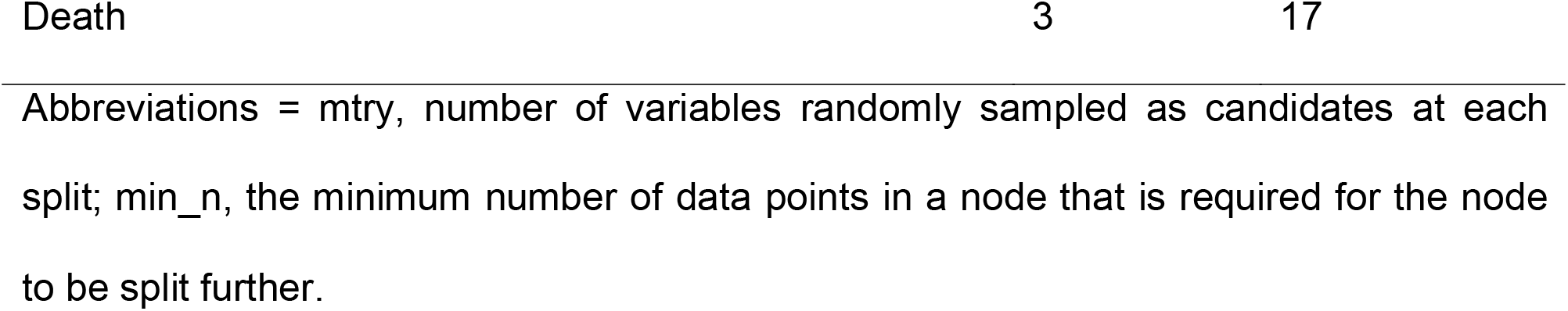
Hyperparameter tuning results.

