## Supplementary Appendix for "Predicting severe COVID-19 outcomes for triage and resource allocation"

### Supporting information

**S1 Fig. Forest plot for mechanical ventilation logistic regression model with all variables.** Male is the reference group for sex. Age 20-29 Years is the reference group for age. Non-Hispanic White is the reference group for race/ethnicity. Northeast is the reference group for the region. The reference group for the month of the positive test is Jan-March.

**S2 Fig. Forest plot for pneumonia logistic regression model with all variables.** Male is the reference group for sex. Age 20-29 Years is the reference group for age. Non-Hispanic White is the reference group for race/ethnicity. Northeast is the reference group for the region. The reference group for the month of the positive test is Jan-March.

**S3 Fig. Forest plot for acute respiratory distress logistic regression model – all variables.** Male is the reference group for sex. Age 20-29 Years is the reference group for age. Non-Hispanic White is the reference group for race/ethnicity. Northeast is the reference group for the region. The reference group for the month of the positive test is Jan-March.

**S1 Table. Model fit statistics for hospitalization logistic regression model.**

| Metric | Full Model with All Variables |
| --- | --- |
| AIC | 921,131.2 |
| BIC | 921,589.4 |
| AUC | 0.121 |
| McFadden’s R2 | 0.341 |
| Nagelkerke’s R2 | 0.433 |

Abbreviations = AIC, Akaike information criterion; BIC, Bayesian information criterion; AUC; area under the curve.

**S2 Table. Model fit statistics for ICU admission logistic regression models.**

| Metric | Full Model with All Variables |
| --- | --- |
| AIC | 237,255.7 |
| BIC | 237,684.0 |
| AUC | 0.139 |
| McFadden’s R^2^ | 0.238 |
| Nagelkerke’s R^2^ | 0.275 |

Abbreviations = AIC, Akaike information criterion; BIC, Bayesian information criterion; AUC; area under the curve.

**S3 Table. Model fit statistics for mechanical ventilation logistic regression models.**

| Metric | Full Model with All Variables | Model with LASSO Selected Variables |
| --- | --- | --- |
| AIC | 112,975.3 | 118,124.7 |
| BIC | 113,393.8 | 118,520.6 |
| AUC | 0.103 | 0.118 |
| McFadden’s R2 | 0.310 | 0.278 |
| Nagelkerke’s R2 | 0.339 | 0.306 |

Abbreviations = AIC, Akaike information criterion; BIC, Bayesian information criterion; AUC; area under the curve; LASSO, least absolute shrinkage and selection operator.

**S4 Table. Model fit statistics for pneumonia logistic regression models.**

| Metric | Full Model with All Variables | Model with LASSO Selected Variables |
| --- | --- | --- |
| AIC | 294,942.5 | 295,800.1 |
| BIC | 295,371.1 | 296,205.5 |
| AUC | 0.167 | 0.169 |
| McFadden’s R^2^ | 0.214 | 0.212 |
| Nagelkerke’s R^2^ | 0.255 | 0.253 |

Abbreviations = AIC, Akaike information criterion; BIC, Bayesian information criterion; AUC; area under the curve; LASSO, least absolute shrinkage and selection operator.

**S5 Table. Model fit statistics for acute respiratory distress logistic regression models.**

| Metric | Full Model with All Variables | Model with LASSO Selected Variables |
| --- | --- | --- |
| AIC | 115,744.8 | 116,143.1 |
| BIC | 116,173.3 | 116,548.5 |
| AUC | 0.164 | 0.167 |
| McFadden’s R^2^ | 0.180 | 0.178 |
| Nagelkerke’s R^2^ | 0.194 | 0.191 |

Abbreviations = AIC, Akaike information criterion; BIC, Bayesian information criterion; AUC; area under the curve; LASSO, least absolute shrinkage and selection operator.

**S6 Table. Model fit statistics for mortality logistic regression model.**

| Metric | Full Model with All Variables |
| --- | --- |
| AIC | 346,641.7 |
| BIC | 347,098.9 |
| AUC | 0.048 |
| McFadden’s R^2^ | 0.483 |
| Nagelkerke’s R^2^ | 0.531 |

Abbreviations = AIC, Akaike information criterion; BIC, Bayesian information criterion; AUC; area under the curve; LASSO, least absolute shrinkage and selection operator.

**S7 Table. Hyperparameter tuning results.**

| Outcome | mtry | min_n |
| --- | --- | --- |
| Hospitalization | 3 | 17 |
| ICU Admission | 3 | 17 |
| Mechanical Ventilation | 3 | 17 |
| Pneumonia | 1 | 15 |
| Acute Respiratory Distress | 1 | 20 |
| Death | 3 | 17 |

Abbreviations = mtry, number of variables randomly sampled as candidates at each split; min_n, the minimum number of data points in a node that is required for the node to be split further.
