## Supplementary material for "Predicting severe COVID-19 outcomes for triage and resource allocation": Letter of Ethical Approval

GUTHRIE

**Institutional Review Board of The Guthrie Clinic**

Michael Georgetson, MD, FACG
*IRB Chair*
Donald Guthrie Foundation
One Guthrie Square
Sayre, PA  18840
570-887-4885


April 12, 2021

Dr. Burt Cagir and Anthony Morada,

Your research project *Predicting severe COVID-19 outcomes for triage and resource allocation* is not considered human subjects research.

Collection of the original data is in compliance with the Office of Human Research Protections (OHRP) and the policies of the Institutional Review Board of The Guthrie Clinic.

Data used in the study will not contain individually identifiable data. The data is a limited data set, and a data use agreement was completed. Access to the data was granted after registration and completion of the *CDC COVID-19 Case Surveillance Restricted Access Detailed Data Registration Information and Data Use Restrictions Agreement (RIDURA)*

Approval by the IRB is not required for use of the limited data set. OHRP does not ordinarily consider such information to be individually identifiable to the investigator if the investigator and the holder of the individually identifying information sign an agreement prohibiting the release of individually identifying information to the investigator under any circumstances.

Please contact me at (570) 887-4885 or if you have any questions.

Sincerely,

Michael Georgetson, MD, FACG
Chairman, Institutional Review Board

Association for the Accreditation of Human Research Protection Programs, Inc.
Full Accreditation
